# Impact of COVID-19 and effects of booster vaccination with BNT162b2 on six-month long COVID symptoms, quality of life, work productivity and activity impairment during Omicron

**DOI:** 10.1101/2023.03.15.23286981

**Authors:** Manuela Di Fusco, Xiaowu Sun, Mary M. Moran, Henriette Coetzer, Joann M. Zamparo, Mary B. Alvarez, Laura Puzniak, Ying P. Tabak, Joseph C. Cappelleri

## Abstract

**Background:** Longitudinal estimates of long COVID burden during Omicron remain limited. This study characterized long-term impacts of COVID-19 and booster vaccination on symptoms, Health-Related Quality of Life (HRQoL), and Work Productivity Activity Impairment (WPAI).

**Methods:** Outpatients with ≥1 self-reported symptom and positive SARS-CoV-2 test at CVS Health United States test sites were recruited between 01/31-04/30/2022. Symptoms, EQ-5D and WPAI were collected via online surveys until 6 months following infection. Both observed and model-based estimates were analyzed. Effect sizes based on Cohen’s d quantified the magnitude of outcome changes over time, within and between vaccination groups. Mixed models for repeated measures were conducted for multivariable analyses, adjusting for covariates. Logistic regression assessed odds ratio (OR) of long COVID between vaccination groups.

**Results:** At long COVID start (Week 4), 328 participants included 87 (27%) Boosted with BNT162b2, 86 (26%) with a BNT162b2 primary series (Primed), and 155 (47%) Unvaccinated. Mean age was 42.0 years, 73.8% were female, 26.5% had ≥1 comorbidity, 36.9% prior infection, and 39.6% reported ≥3 symptoms (mean: 3.1 symptoms). At Month 6, among 260 participants, Boosted reported a mean of 1.1 symptoms versus 3.4 and 2.8 in Unvaccinated and Primed, respectively (p<0.001). Boosted had reduced risks of ≥3 symptoms versus Unvaccinated (observed: OR 0.22, 95% CI, 0.10-0.47, p<0.001; model-based: OR: 0.36, 95% CI, 0.15-0.87, p=0.019) and Primed (observed: OR 0.29, 95% CI, 0.13-0.67, p=0.003; model-based: OR 0.59, 95% CI, 0.21-1.65, p=0.459). Results were consistent using ≥2 symptoms. Regarding HRQoL, among those with long COVID, Boosted had higher EQ-5D Utility Index (UI) than Unvaccinated (observed: 0.922 versus 0.731, p=0.014; model-based: 0.910 versus 0.758, p-value=0.038) and Primed (0.922 versus 0.648, p=0.014; model-based: 0.910 versus 0.708, p-value=0.008). Observed and model-based estimates for EQ-VAS and UI among Boosted were comparable with pre-COVID since Month 3. Subjects vaccinated generally reported better WPAI scores.

**Conclusions:** Long COVID negatively impacted HRQoL and WPAI. The BNT162b2 booster could have a beneficial effect in reducing the risk and burden of long COVID. Boosted participants reported fewer and less durable symptoms, which contributed to improve HRQoL and maintain WPAI levels. Limitations included self-reported data and small sample size for WPAI.

## Background

Long COVID is a broad array of complications of SARS-CoV-2 infection [1]. There is no single definition of long-COVID presently and prevalence estimates vary widely across studies and variants [1]. In the US, the Centers for Disease Control and Prevention (CDC) define long COVID as a multitude of symptoms and conditions persisting or emerging beyond four weeks after infection, and not explained by an alternative diagnosis [1]. Long COVID is found more often in people who had severe COVID-19 illness, but anyone who was infected can experience it [2]. While most individuals with long COVID are mildly affected and recover shortly after infection, some can suffer from persistent symptoms that can affect the Health-Related Quality of Life (HRQoL) and impact daily social and professional activities [3].

Multiple studies assessed whether vaccinated individuals are associated with a lower risk of long COVID, several of them show that long COVID is less likely to occur following breakthrough infection in vaccinated compared with unvaccinated individuals [4–7]. However, numerous studies were conducted in selected populations that may affect generalizability [4, 5, 7, 8], assessed a mix of COVID-19 vaccines, and the certainty of evidence was considered low when pooling studies [6, 8]. Moreover, there is limited data on the long-term effects of COVID-19 and the potential benefits of COVID-19 vaccination on symptoms, well-being, and ability to return to work.

We aimed to address these gaps using a prospective longitudinal study design assessing long COVID prevalence and levels of HRQoL and WPAI through 6 months following infection, and among groups defined by COVID-19 vaccination status. Our study focused on BNT162b2, the most used COVID-19 vaccine in the US [1].

## Methods

### Study design and cohorts

The study design was previously described [9] (clinicatrials.gov NCT05160636). Briefly, this was a nationwide prospective survey-based patient-reported outcomes (PRO) study targeting adults ≥18 years old with a positive reverse transcription–polymerase chain reaction (RT-PCR) test result and self-reporting at least one symptom suggestive of acute SARS-CoV-2 infection at time of testing [9]. The source population consisted of adults testing for SARS-CoV-2 at one of ∼5,000 CVS Health test sites across the US. Recruitment of consenting participants was carried out between 01/31/2022 and 04/30/2022, with follow-up occurring through October 30, 2022. Interim results for the acute phase (up to 4 weeks after infection) were previously presented [9]. This analysis presents the final results, focusing on long-term outcomes until month 6.

At enrollment, we categorized the participants based on their COVID-19 vaccination history. Immunocompetent participants were considered fully vaccinated with BNT162b2 if they self-reported receipt of 2 doses of BNT162b2 ≥ 14 days prior to SARS-CoV-2 testing. They were considered partially vaccinated if reporting receipt of only one of the two primary series dose, and boosted if reporting receipt of at least one dose after the primary series. Participants self-reporting an immunocompromising condition and receipt of 3 doses were considered fully vaccinated; if reporting 4 doses, they were considered boosted. Participants were considered unvaccinated if they did not report any COVID-19 vaccine dose prior to testing. The study population was classified in three mutually exclusive cohorts: (1) the “Primed” cohort of subjects who received primary series, (2) the “Boosted” cohort of subjects with at least one dose after primary series, and (3) the “Unvaccinated” cohort of subject with no evidence of vaccination. Heterologous schedules were excluded (Supplemental Figure 1). To confirm vaccination status, participants’ subsequent responses to vaccination date questions were compared with their index responses (at time of registering for testing); if responses did not match, the information was queried and adjudicated, and the latest information was used.

### Baseline characteristics and symptoms

Baseline characteristics were obtained via the CVS Health pre-test screening questionnaire. These included self-reported demographics, comorbidities, COVID-19 vaccination history, social determinants of health including the Social Vulnerability Index (SVI), work and/or residency in a high-risk or healthcare setting, and symptoms [10]. The list of COVID-19 symptoms came from the CDC’s definition [11].

### Long COVID Symptoms

The presence of long COVID symptoms was assessed via a questionnaire including 20 symptoms based on the 2021 CDC list [1]. The questionnaire was administered at week 4, month 3 and 6 after enrollment. Week 4 was considered the start of long COVID, in alignment with CDC [1]. The list of symptoms included general symptoms (tiredness or fatigue, symptoms that get worse after physical or mental activities, fever, general pain/discomfort), respiratory and cardiac symptoms (difficulty breathing or shortness of breath, cough, chest or stomach pain, fast-beating or pounding heart (also known as heart palpitations)), neurological symptoms (change in smell or taste, headache, dizziness on standing (lightheadedness), difficulty thinking or concentrating ( “brain fog”), pins-and-needles feeling, sleep problems, mood changes, memory loss), and other symptoms (diarrhea, joint or muscle pain, rash, changes in period cycles). (Supplemental Figure 2).

Studies describing the prevalence of long COVID in the general population testing positive for COVID-19 have used different thresholds for duration and intensity of symptoms [12–14]. Interim data from the CDC-funded INSPIRE Registry reported long COVID among subjects who were symptomatic at time of testing using a cutoff of ≥3 symptoms at 3 months post infection, with follow-up surveys scheduled every three months until 18 months post-enrollment [15, 16]. Considering the similarities in the total number of symptoms assessed (∼20), eligibility criteria and study design, this study used a similar cut-off of ≥3 symptoms, assessed 3- and 6-months following infection

### Health-Related Quality of Life

We used EQ-5D-5L to assess HRQoL [17]. Completion of the questionnaire was requested at enrollment, at month 1, 3 and 6 [9]. Five dimensions of EQ-5D-5L at each time point were converted into the Utility Index (UI) using the US-based weights by Pickard et al [18]. UI and visual analogue scale (VAS) scores were compared among cohorts and across assessment times [17].

### Work Productivity and Activity Impairment

To measure impairments in both paid work and unpaid work, we used the Work Productivity and Activity Impairment General Health V2.0 (WPAI:GH) measure [19, 20]. Participants were asked to complete the survey seven days after their RT-PCR test, at month 1, 3 and 6. Only employed subjects were included for work productivity analyses. The WPAI results were compared across cohorts and assessment times.

### Statistical methods

The statistical methods were previously described [9]. Descriptive statistics were used to analyze participant characteristics at baseline. Continuous variables were described using means and standard deviations. Categorical variables were summarized with frequency and percentages. For continuous variables, t-tests were used to test difference in means. Between-group differences in categorical variables were tested by using chi-square statistics. When expected cell frequency count was less than 5, Fisher’s exact tests were used for 2-by-2 tables and Freeman-Halton tests for r-by-c tables [21, 22]. Analysis of variance (ANOVA) was used to test differences in means. Tukey’s studentized range test was adopted for post ANOVA pairwise comparisons of means between study cohorts [21, 23]. P values were all two-sided. Logistic regression model [20] was used to assess the odds ratio of long COVID symptoms between study cohorts.

Mixed models for repeated measures (MMRM) were used to estimate the magnitude of COVID-19 impact on HRQoL and WPAI over time [24]. Assessment time was fitted as a categorical covariate and a repeated effect (repeated by subject). Least squares mean (LS mean) and standard errors of PRO scores for each time point of assessment were calculated. Per guidelines, no adjustment was made for missing data when scoring the EQ-5D-5L UI and WPAI [17, 20]. All available data were included in the analysis. Tukey’s adjustment was conducted for the comparisons of least-square means between study cohorts at each time point.

Cohen’s d, or a variation of it, was calculated to assess the difference in pre-COVID scores among Boosted, Primed and Unvaccinated, the magnitude of score change from baseline at Week 4, Month 3 and 6 within each cohort, as well as the differences between cohorts [25, 26]. Specifically, within-cohort effect size (ES) was calculated as mean change from baseline to follow-up, divided by the standard deviation of change scores from baseline to follow-up [9]. Between-cohort ES was calculated as the difference in mean score between cohorts, divided by the pooled standard deviation of scores, or the difference in mean changes from baseline between cohorts, divided by the pooled standard deviation of change scores [9]. Values of 0.2, 0.5, and 0.8 standard deviation (SD) units were taken represent “small,” “medium,” and “large” effect sizes, respectively [9, 25]. All analyses were conducted with SAS Version 9.4 (SAS Institute, Cary, NC). The study followed the Strengthening the Reporting of Observational Studies in Epidemiology (STROBE) reporting guideline [27].

## Results

### Study population

Table 1 describes the characteristics of the 328 participants at baseline (i.e., four weeks after infection). There were 87 (27%) Boosted, 86 (26%) Primed, and 155 (47%) Unvaccinated. There were no subjects partially vaccinated. At Month 6, 21% (68/328) of participants dropped off. Mean time from last dose of BNT162b2 to testing positive was 2.3 (standard deviation, SD: 1.9) months and 6.9 (SD: 3.0) months for Boosted and Primed, respectively. The population was 42.0 years old on average, 73.8% were female, 26.5% reported ≥1 comorbidity and 36.9% were previously infected.

**Table 1.**
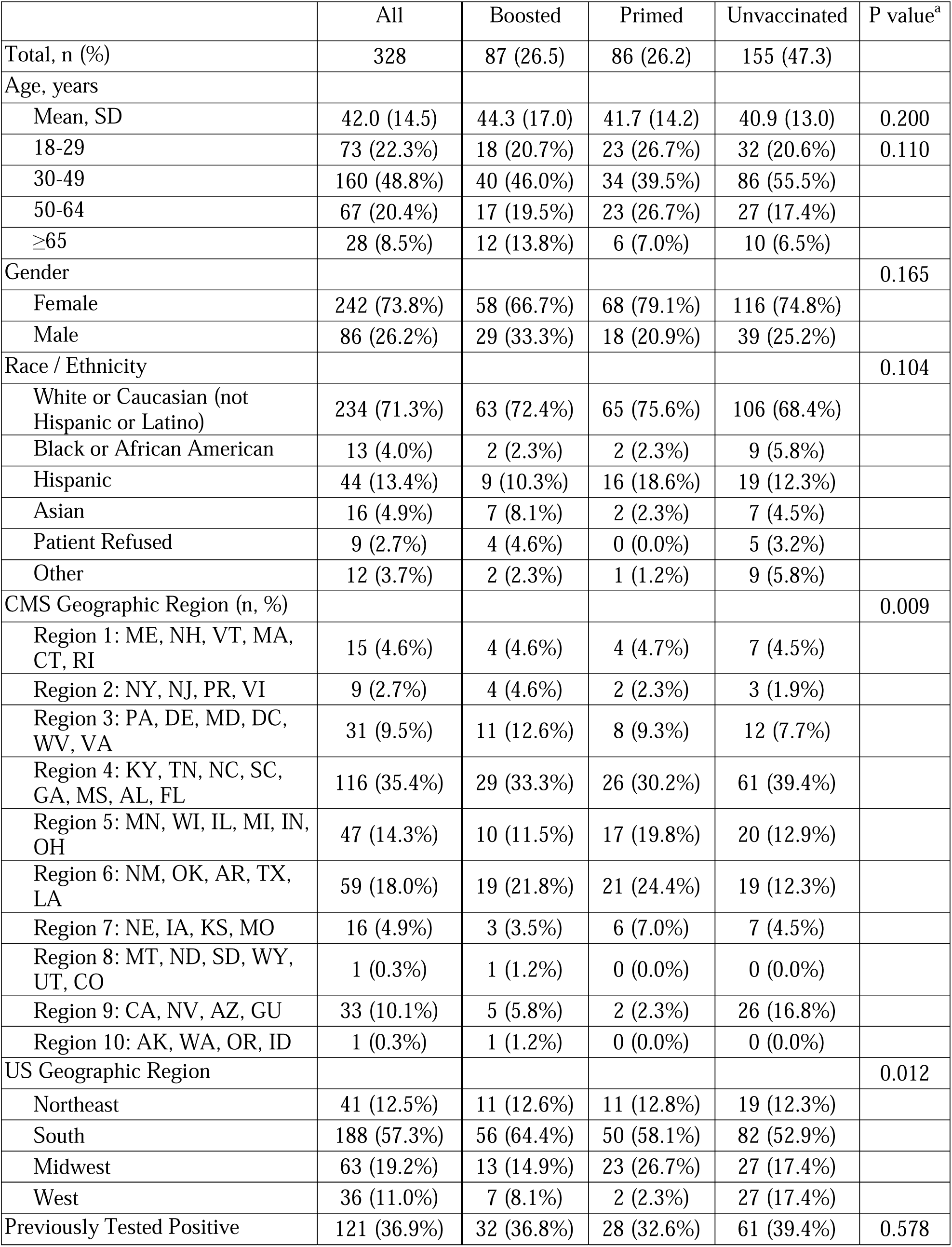
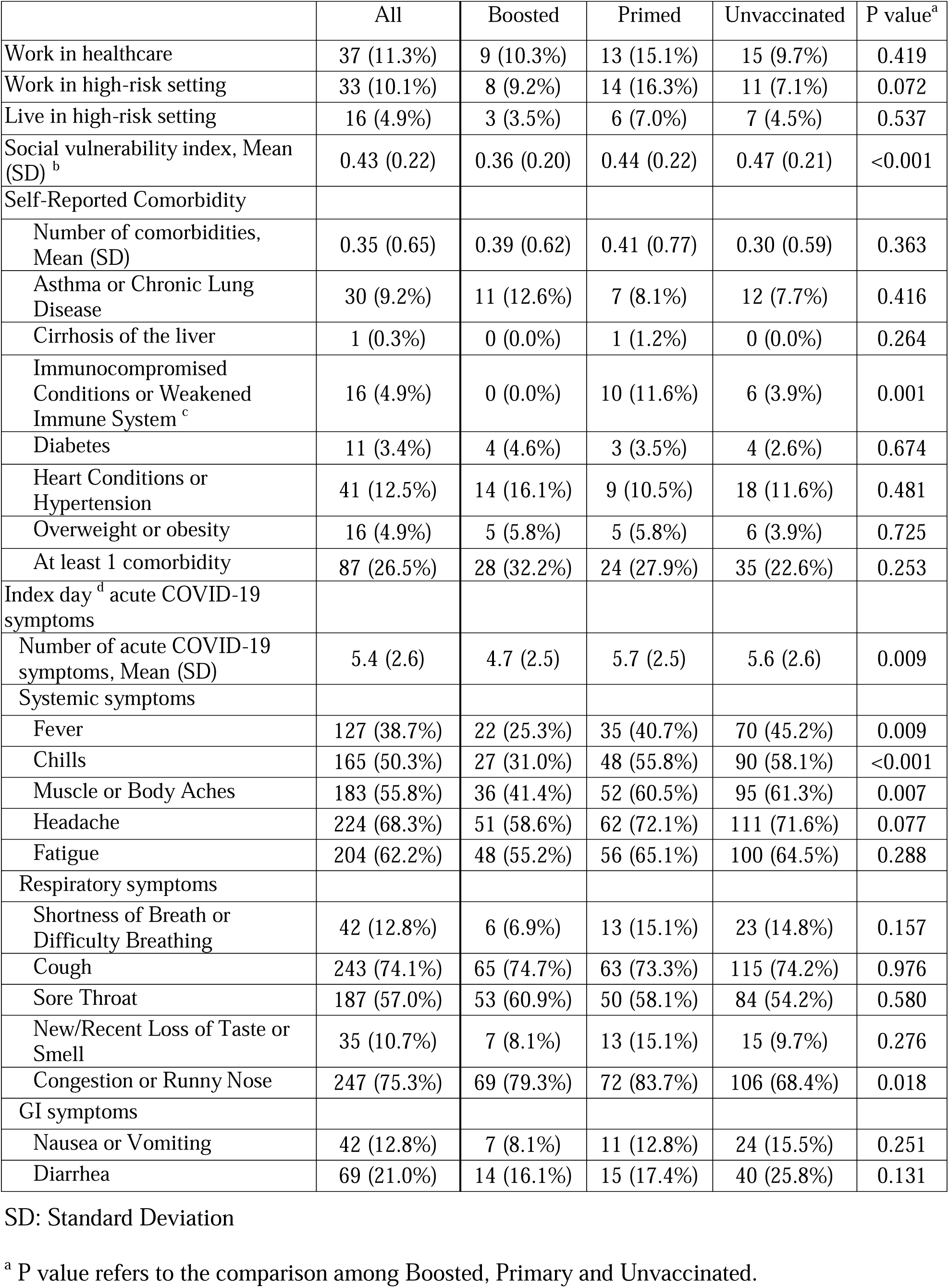
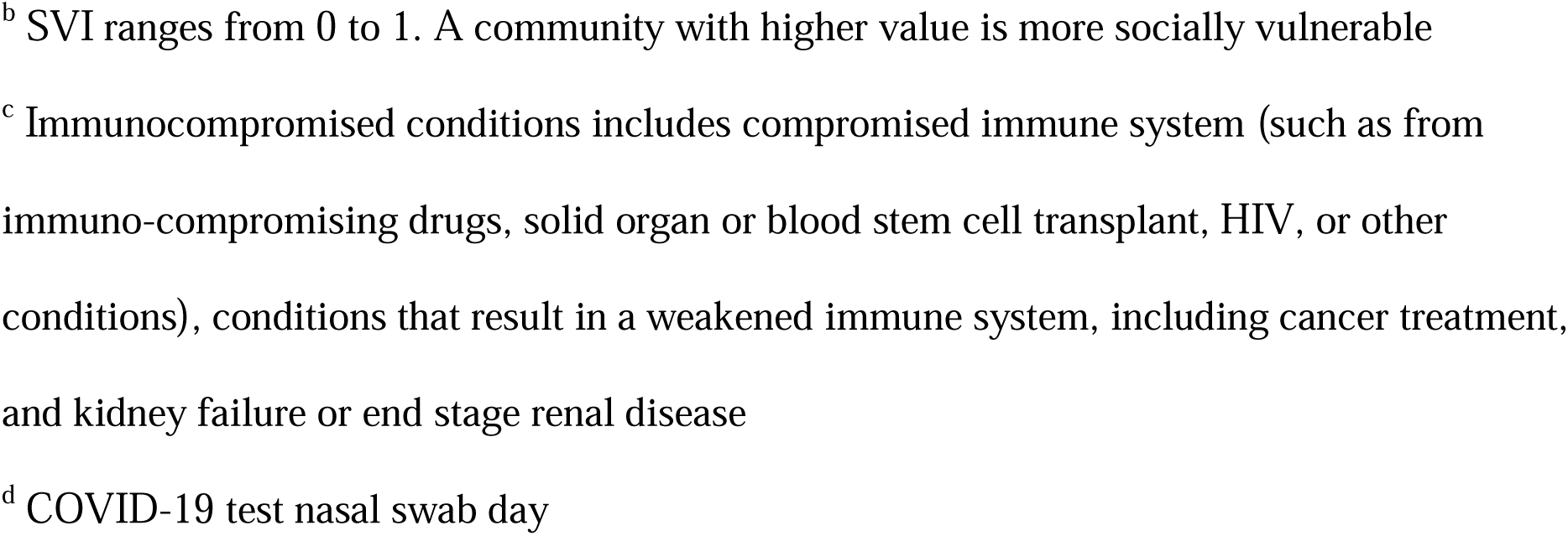
Baseline Demographic and Clinical Characteristics of Study Participants at Week 4

|  | All | Boosted | Primed | Unvaccinated | P value<sup>a</sup> |
| --- | --- | --- | --- | --- | --- |
| Total, n (%) | 328 | 87 (26.5) | 86 (26.2) | 155 (47.3) |  |
| Age, years |  |  |  |  |  |
| Mean, SD | 42.0 (14.5) | 44.3 (17.0) | 41.7 (14.2) | 40.9 (13.0) | 0.200 |
| 18-29 | 73 (22.3%) | 18 (20.7%) | 23 (26.7%) | 32 (20.6%) | 0.110 |
| 30-49 | 160 (48.8%) | 40 (46.0%) | 34 (39.5%) | 86 (55.5%) |  |
| 50-64 | 67 (20.4%) | 17 (19.5%) | 23 (26.7%) | 27 (17.4%) |  |
| ≥65 | 28 (8.5%) | 12 (13.8%) | 6 (7.0%) | 10 (6.5%) |  |
| Gender |  |  |  |  | 0.165 |
| Female | 242 (73.8%) | 58 (66.7%) | 68 (79.1%) | 116 (74.8%) |  |
| Male | 86 (26.2%) | 29 (33.3%) | 18 (20.9%) | 39 (25.2%) |  |
| Race / Ethnicity |  |  |  |  | 0.104 |
| White or Caucasian (not Hispanic or Latino) | 234 (71.3%) | 63 (72.4%) | 65 (75.6%) | 106 (68.4%) |  |
| Black or African American | 13 (4.0%) | 2 (2.3%) | 2 (2.3%) | 9 (5.8%) |  |
| Hispanic | 44 (13.4%) | 9 (10.3%) | 16 (18.6%) | 19 (12.3%) |  |
| Asian | 16 (4.9%) | 7 (8.1%) | 2 (2.3%) | 7 (4.5%) |  |
| Patient Refused | 9 (2.7%) | 4 (4.6%) | 0 (0.0%) | 5 (3.2%) |  |
| Other | 12 (3.7%) | 2 (2.3%) | 1 (1.2%) | 9 (5.8%) |  |
| CMS Geographic Region (n, %) |  |  |  |  | 0.009 |
| Region 1: ME, NH, VT, MA, CT, RI | 15 (4.6%) | 4 (4.6%) | 4 (4.7%) | 7 (4.5%) |  |
| Region 2: NY, NJ, PR, VI | 9 (2.7%) | 4 (4.6%) | 2 (2.3%) | 3 (1.9%) |  |
| Region 3: PA, DE, MD, DC, WV, VA | 31 (9.5%) | 11 (12.6%) | 8 (9.3%) | 12 (7.7%) |  |
| Region 4: KY, TN, NC, SC, GA, MS, AL, FL | 116 (35.4%) | 29 (33.3%) | 26 (30.2%) | 61 (39.4%) |  |
| Region 5: MN, WI, IL, MI, IN, OH | 47 (14.3%) | 10 (11.5%) | 17 (19.8%) | 20 (12.9%) |  |
| Region 6: NM, OK, AR, TX, LA | 59 (18.0%) | 19 (21.8%) | 21 (24.4%) | 19 (12.3%) |  |
| Region 7: NE, IA, KS, MO | 16 (4.9%) | 3 (3.5%) | 6 (7.0%) | 7 (4.5%) |  |
| Region 8: MT, ND, SD, WY, UT, CO | 1 (0.3%) | 1 (1.2%) | 0 (0.0%) | 0 (0.0%) |  |
| Region 9: CA, NV, AZ, GU | 33 (10.1%) | 5 (5.8%) | 2 (2.3%) | 26 (16.8%) |  |
| Region 10: AK, WA, OR, ID | 1 (0.3%) | 1 (1.2%) | 0 (0.0%) | 0 (0.0%) |  |
| US Geographic Region |  |  |  |  | 0.012 |
| Northeast | 41 (12.5%) | 11 (12.6%) | 11 (12.8%) | 19 (12.3%) |  |
| South | 188 (57.3%) | 56 (64.4%) | 50 (58.1%) | 82 (52.9%) |  |
| Midwest | 63 (19.2%) | 13 (14.9%) | 23 (26.7%) | 27 (17.4%) |  |
| West | 36 (11.0%) | 7 (8.1%) | 2 (2.3%) | 27 (17.4%) |  |
| Previously Tested Positive | 121 (36.9%) | 32 (36.8%) | 28 (32.6%) | 61 (39.4%) | 0.578 |
| Work in healthcare | 37 (11.3%) | 9 (10.3%) | 13 (15.1%) | 15 (9.7%) | 0.419 |
| Work in high-risk setting | 33 (10.1%) | 8 (9.2%) | 14 (16.3%) | 11 (7.1%) | 0.072 |
| Live in high-risk setting | 16 (4.9%) | 3 (3.5%) | 6 (7.0%) | 7 (4.5%) | 0.537 |
| Social vulnerability index, Mean (SD)<sup>b</sup> | 0.43 (0.22) | 0.36 (0.20) | 0.44 (0.22) | 0.47 (0.21) | <0.001 |
| Self-Reported Comorbidity |  |  |  |  |  |
| Number of comorbidities, Mean (SD) | 0.35 (0.65) | 0.39 (0.62) | 0.41 (0.77) | 0.30 (0.59) | 0.363 |
| Asthma or Chronic Lung Disease | 30 (9.2%) | 11 (12.6%) | 7 (8.1%) | 12 (7.7%) | 0.416 |
| Cirrhosis of the liver | 1 (0.3%) | 0 (0.0%) | 1 (1.2%) | 0 (0.0%) | 0.264 |
| Immunocompromised Conditions or Weakened Immune System<sup>c</sup> | 16 (4.9%) | 0 (0.0%) | 10 (11.6%) | 6 (3.9%) | 0.001 |
| Diabetes | 11 (3.4%) | 4 (4.6%) | 3 (3.5%) | 4 (2.6%) | 0.674 |
| Heart Conditions or Hypertension | 41 (12.5%) | 14 (16.1%) | 9 (10.5%) | 18 (11.6%) | 0.481 |
| Overweight or obesity | 16 (4.9%) | 5 (5.8%) | 5 (5.8%) | 6 (3.9%) | 0.725 |
| At least 1 comorbidity | 87 (26.5%) | 28 (32.2%) | 24 (27.9%) | 35 (22.6%) | 0.253 |
| Index day<sup>d</sup> acute COVID-19 symptoms |  |  |  |  |  |
| Number of acute COVID-19 symptoms, Mean (SD) | 5.4 (2.6) | 4.7 (2.5) | 5.7 (2.5) | 5.6 (2.6) | 0.009 |
| Systemic symptoms |  |  |  |  |  |
| Fever | 127 (38.7%) | 22 (25.3%) | 35 (40.7%) | 70 (45.2%) | 0.009 |
| Chills | 165 (50.3%) | 27 (31.0%) | 48 (55.8%) | 90 (58.1%) | <0.001 |
| Muscle or Body Aches | 183 (55.8%) | 36 (41.4%) | 52 (60.5%) | 95 (61.3%) | 0.007 |
| Headache | 224 (68.3%) | 51 (58.6%) | 62 (72.1%) | 111 (71.6%) | 0.077 |
| Fatigue | 204 (62.2%) | 48 (55.2%) | 56 (65.1%) | 100 (64.5%) | 0.288 |
| Respiratory symptoms |  |  |  |  |  |
| Shortness of Breath or Difficulty Breathing | 42 (12.8%) | 6 (6.9%) | 13 (15.1%) | 23 (14.8%) | 0.157 |
| Cough | 243 (74.1%) | 65 (74.7%) | 63 (73.3%) | 115 (74.2%) | 0.976 |
| Sore Throat | 187 (57.0%) | 53 (60.9%) | 50 (58.1%) | 84 (54.2%) | 0.580 |
| New/Recent Loss of Taste or Smell | 35 (10.7%) | 7 (8.1%) | 13 (15.1%) | 15 (9.7%) | 0.276 |
| Congestion or Runny Nose | 247 (75.3%) | 69 (79.3%) | 72 (83.7%) | 106 (68.4%) | 0.018 |
| GI symptoms |  |  |  |  |  |
| Nausea or Vomiting | 42 (12.8%) | 7 (8.1%) | 11 (12.8%) | 24 (15.5%) | 0.251 |
| Diarrhea | 69 (21.0%) | 14 (16.1%) | 15 (17.4%) | 40 (25.8%) | 0.131 |

SD: Standard Deviation
<sup>a</sup> P value refers to the comparison among Boosted, Primary and Unvaccinated.
<sup>b</sup> SVI ranges from 0 to 1. A community with higher value is more socially vulnerable <sup>c</sup> Immunocompromised conditions includes compromised immune system (such as from immuno-compromising drugs, solid organ or blood stem cell transplant, HIV, or other conditions), conditions that result in a weakened immune system, including cancer treatment, and kidney failure or end stage renal disease <sup>d</sup> COVID-19 test nasal swab day

The three groups generally looked similar in terms of age, gender, race, prior infection, number of comorbidities and risk levels in workplace and household settings. Boosted participants were associated with less social vulnerability, with a mean SVI of 0.36 compared with 0.44 in Primed and 0.47 in Unvaccinated (p<0.001). Primed and Unvaccinated looked similar in terms of SVI, comorbidities, and acute COVID-19 symptoms. Boosted reported fewer acute COVID-19 symptoms, with a mean of 4.7 acute symptoms compared with 5.7 in Primed and 5.6 in Unvaccinated (p=0.009). Systemic symptoms (fever, chills and muscle or body aches) and congestion/ runny nose among Boosted were significantly lower than Primed and Unvaccinated.

### Long COVID-19 risk and symptoms

#### Time trends through Month 6

Prevalence rates of long COVID-19 by assessment time and vaccination status are presented in Table 2 and Supplemental Figure 3. Overall, 39.6%, 37.3% and 35.0% experienced ≥3 symptoms at Week 4, Month 3 and 6, respectively. The most prevalent symptom was consistently reported to be “Fatigue or Tiredness”, experienced by 41.2%, 37.3% and 38.8% at Week 4, Month 3 and 6, respectively.

**Table 2.**
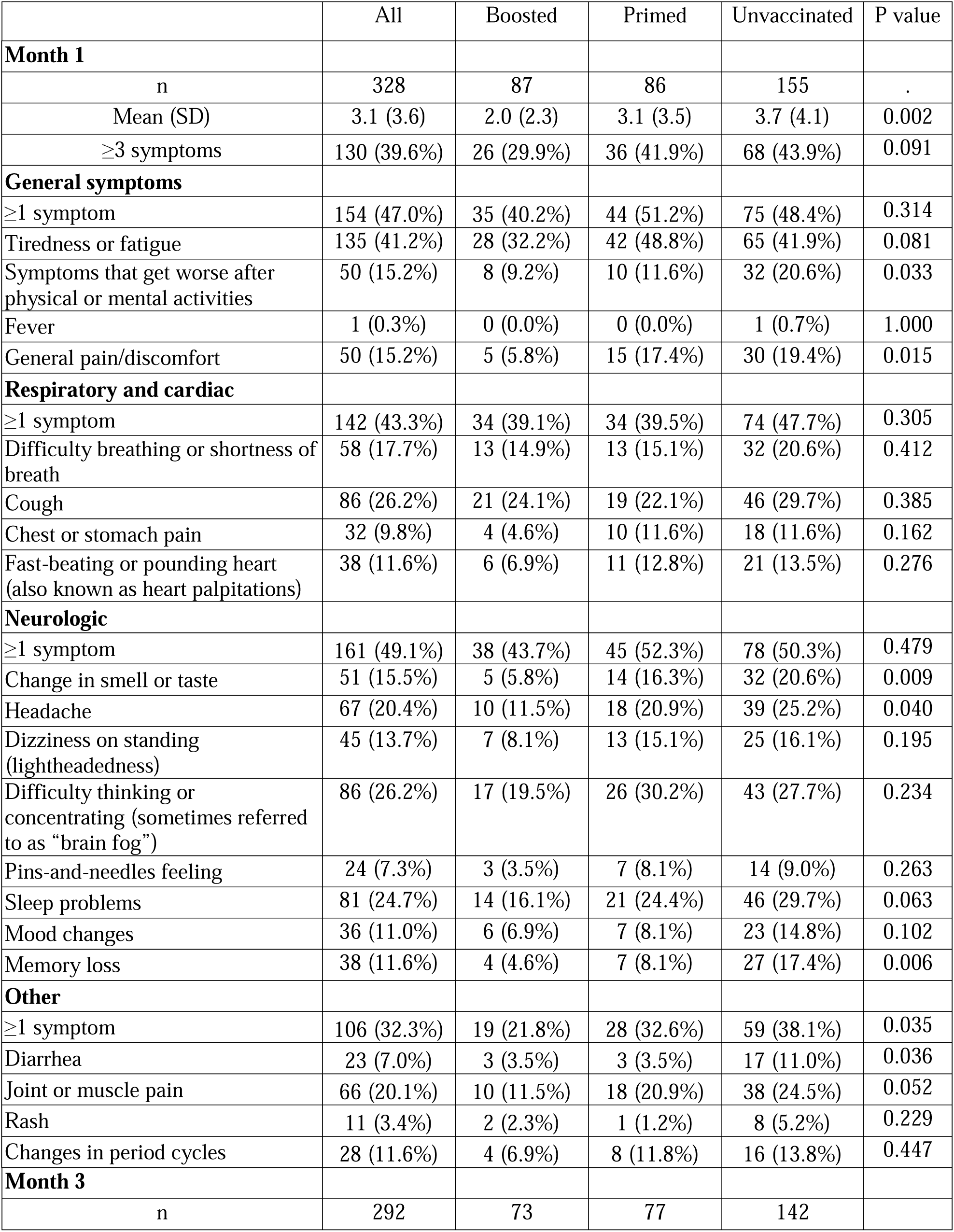
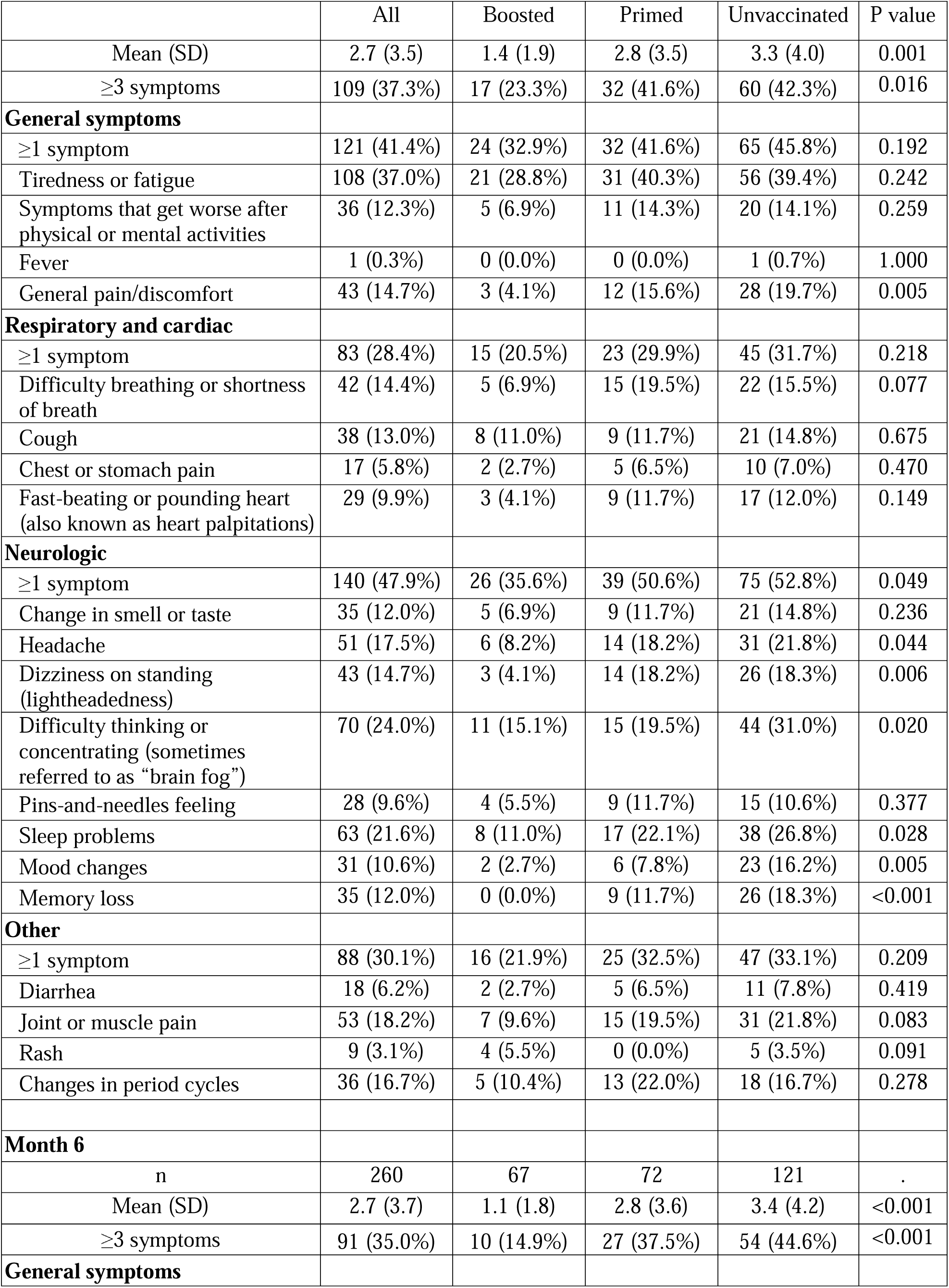
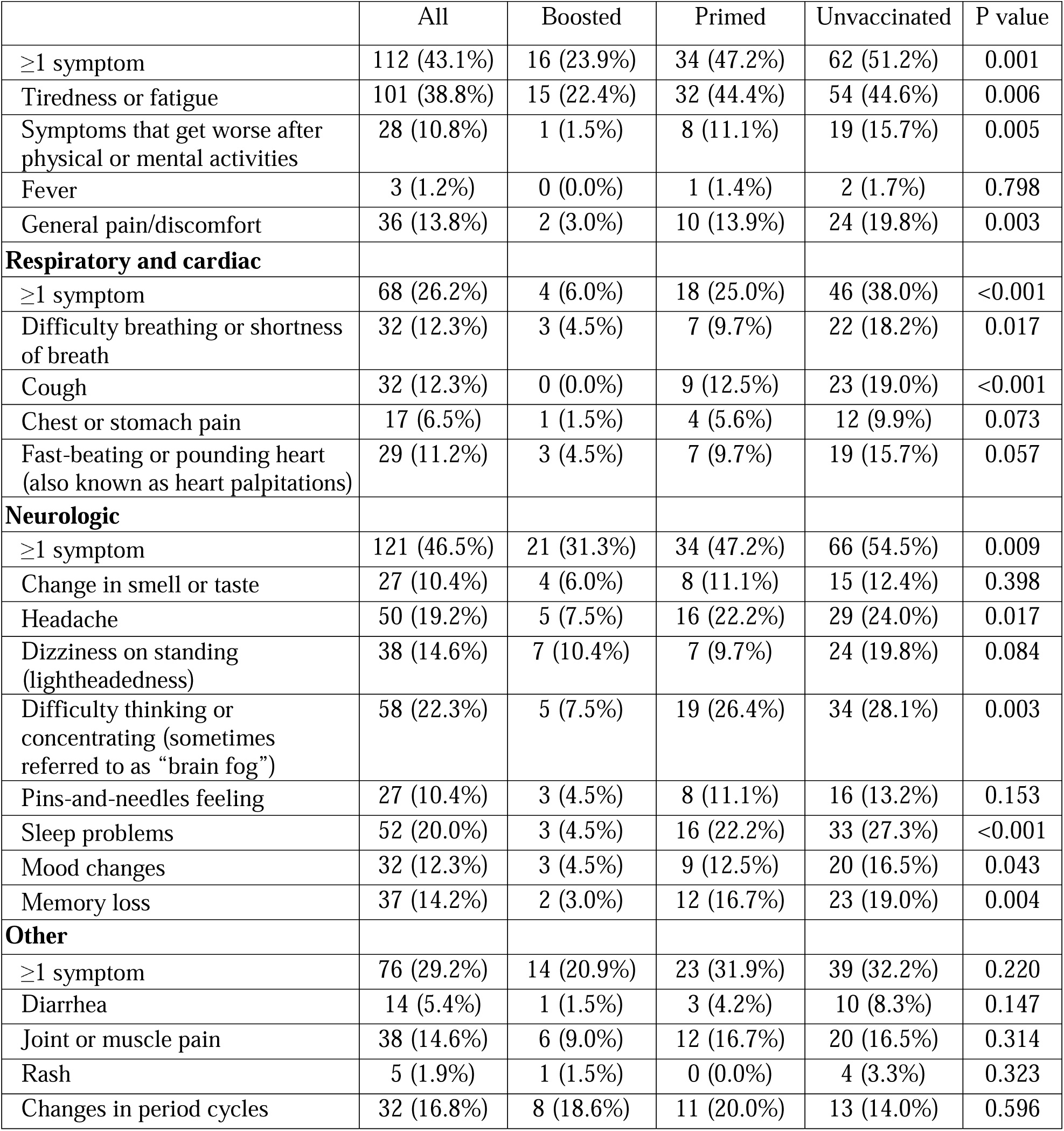
Summary of Post-COVID-19 Symptoms at 1-, 3- and 6-month follow-up

Figure 1 illustrates time trends of prevalence rates of long COVID symptoms by vaccination status. Across all time points, the Boosted line appeared below and separated from the ones of Unvaccinated and Primed when using the base case definition of long COVID with a cut-off of ≥3 symptoms (Figure 1a), as well as when using a cutoff of ≥2 symptoms (Figure 1b). Figures 1c-1f show the prevalence rates of long COVID by body system: across all time points and symptom type, the Boosted line appeared below and separated from the ones of Unvaccinated and Primed. Among Boosted, declines were visible at Month 6 in Respiratory and Cardiac Symptoms, General Symptoms, and Neurologic Symptoms.

**Figure 1.**
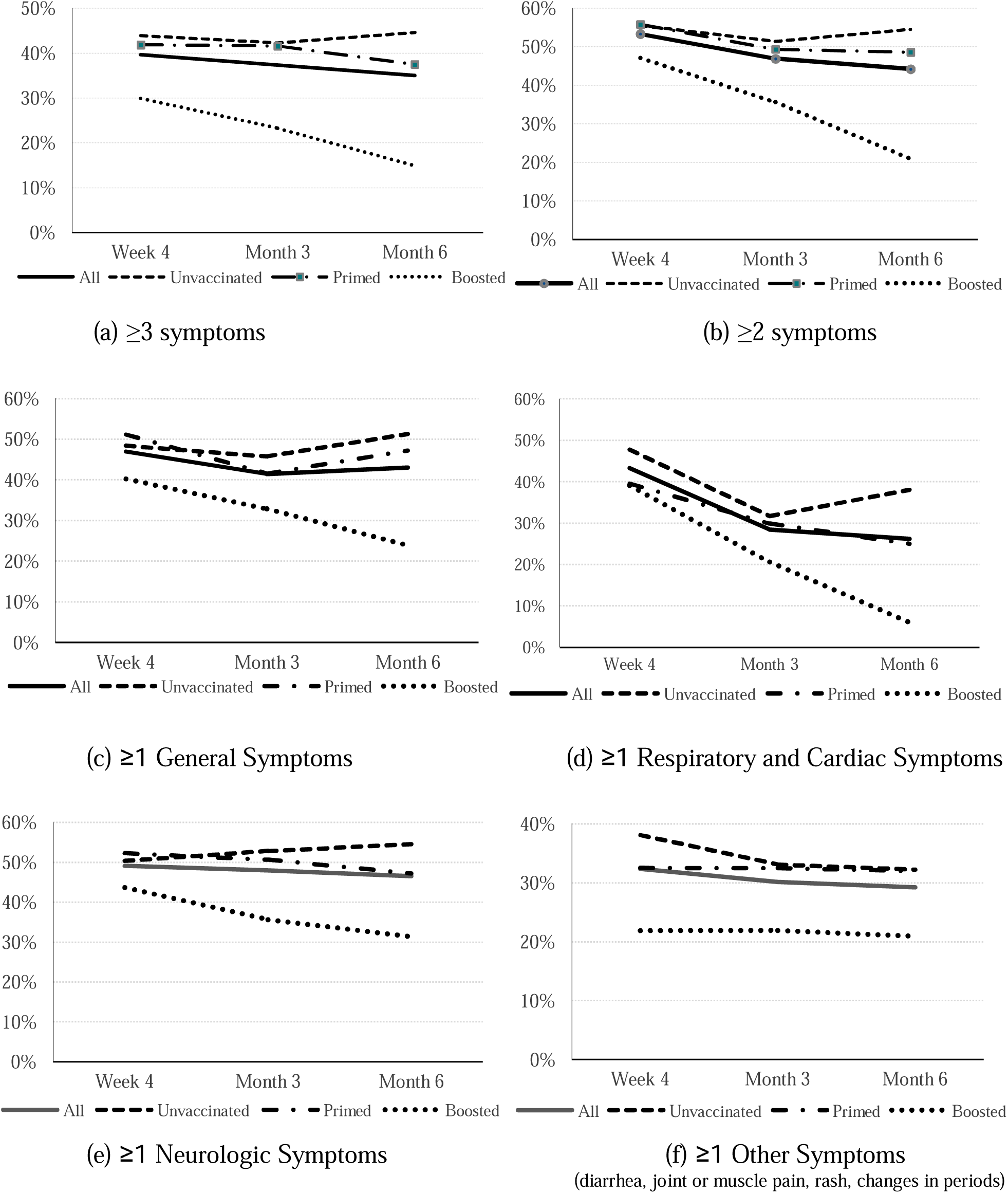
Trajectory of prevalence of long COVID over time, by vaccination status and type

From Week 4 to Month 6, noticeable improvements were observed among Boosted in several symptoms, such as Tiredness or Fatigue (32.2% to 22.4%), Cough (24.1% to 0.0%), Difficulty thinking or concentrating (19.5% to 7.5%), Difficulty breathing or shortness of breath (14.9% to 4.5%), Sleep problems (16.1% to 4.5%), and Symptoms that get worse after physical or mental activities (9.2% to 1.5%). Supplemental Figures 4 and 5 illustrate, respectively, time trends of the mean numbers of symptoms by vaccination status, and the proportions of participants reporting no symptoms over time: the Boosted lines were consistently separated from the ones of Unvaccinated and Primed.

#### Week 4

At long COVID start, participants reported a mean of 3.1 symptoms. Boosted reported fewer symptoms: on average, 2.0 symptoms, significantly lower than 3.1 in Primed and 3.7 in Unvaccinated (p=0.002). The prevalence of long COVID (≥3 symptoms) was 29.9%, 41.9%, and 43.9% among Boosted, Primed and Unvaccinated, respectively, being directionally lower in Boosted (p=0.091). Relative to Unvaccinated, all symptoms’ prevalence rates were numerically lower in Boosted, and numerically lower or similar relative to Primed. Several symptoms were significantly lower: symptoms worsening after physical or mental activities (8.5%), General pain/discomfort (5.8%), Change in smell or taste (5.8%), Headache (11.5%), Memory loss (4.6%), and Diarrhea (3.5%) (Table 2).

#### Month 3

At Month 3, the prevalence of long-COVID was 23.3%, 41.6%, and 42.3% among Boosted, Primed and Unvaccinated, respectively, being lowest in Boosted (p=0.016). Study participants reported a mean of 2.7 symptoms. Subjects Boosted reported on average 1.4 symptoms, significantly lower than 2.8 in Primed and 3.3 in Unvaccinated (p=0.001). (Table 2) All symptoms’ prevalence rates were numerically lower in Boosted compared to Unvaccinated and Primed. Significantly lower rates were reported for Boosted for General pain/discomfort (4.1%), Headache (8.2%), Dizziness on standing (lightheadedness) (4.1%), Difficulty thinking or concentrating (15.1%), Sleep problems (11.0%), Mood changes (2.7%), and Memory loss (0.0%) (Table 2).

#### Month 6

At the end of the study period, the prevalence of long-COVID was 14.9%, 37.5%, and 44.6% among Boosted, Primary and Unvaccinated, respectively, being lowest among Boosted (p<0.001). Odds Ratios (OR) calculated based on observed data showed that pre-COVID booster vaccination was associated with a reduced risk of long COVID (expressed as ≥3 symptoms) versus Unvaccinated (OR=0.22, 95% CI: 0.10-0.47, p<0.001) and Primed (OR=0.29, 95% CI: 0.12-0.67, p=0.003). Those Primed were associated with a non-significant reduction in the odds of long COVID against Unvaccinated (OR=0.74, 95% CI: 0.41-1.35, p=0.332). The logistic regression model provided a similar result of reduced odds of long COVID for Boosted versus Unvaccinated (OR=0.36, 95% CI: 0.15-0.87, p=0.019), and a non-significant reduction in the odds of long COVID against Primed (OR=0.59, 95% CI: 0.21-1.65, p=0.459). Similarly, a non-significant reduction in the odds of long COVID was estimated for Primed versus Unvaccinated (OR=0.60, 95% CI: 0.27-1.34, p=0.296). Results were consistent when using an alternative definition of long COVID as ≥2 symptoms (Table 3).

**Table 3.** Long COVID Symptoms at Month 6 by Vaccination Status: Observed and Model-based Estimates

|  | Descriptive statistics | Observed |  | Logistic Regression |  |
| --- | --- | --- | --- | --- | --- |
|  |  | Odds Ratio (95% CI) | P value | Odds Ratio (95% CI) | P value |
| Base Case: ≥3 symptoms |  |  |  |  |  |
| Boosted vs Unvaccinated | 10 (14.9%) vs 54 (44.6%) | 0.22 (0.10, 0.47) | <0.001 | 0.36 (0.15, 0.87) | 0.019 |
| Primed vs. Unvaccinated | 27 (37.5%) vs 54 (44.6%) | 0.74 (0.41, 1.35) | 0.332 | 0.60 (0.27, 1.34) | 0.296 |
| Boosted vs Primed | 10 (14.9%) vs 27 (37.5%) | 0.29 (0.13, 0.67) | 0.003 | 0.59 (0.21, 1.65) | 0.459 |
| Sensitivity: ≥2 symptoms |  |  |  |  |  |
| Boosted vs Unvaccinated | 14 (20.9%) vs 66 (54.5%) | 0.22 (0.11, 0.44) | <0.001 | 0.30 (0.13, 0.70) | 0.003 |
| Primed vs. Unvaccinated | 35 (48.6%) vs 66 (54.5%) | 0.79 (0.44, 1.41) | 0.425 | 0.64 (0.30, 1.39) | 0.370 |
| Boosted vs Primed | 14 (20.9%) vs 35 (48.6%) | 0.28 (0.13, 0.59) | 0.001 | 0.46 (0.18, 1.20) | 0.140 |

Note for logistic regression model: the logistic regression model for number of post-COVID $\geq 3$ versus $< 3$ used GEE estimation with unstructured correlation matrix. Covariates were variables for time, vaccination status and interaction of time by vaccination status, as well as covariates of participant pre-COVID-19 symptom onset score, sociodemographic characteristics (age, sex, regions, social vulnerability, race/ethnicity, high risk occupations), previously tested positive for COVID-19, severity of acute illness (number of symptoms reported on index date), and immunocompromised status.

Study participants reported a mean of 2.7 symptoms at 6 months. The average number of symptoms was 1.1 in Boosted, significantly lower than 2.8 in Primed and 3.4 in Unvaccinated (p<0.001) (Table 2). All symptoms’ prevalence rates were numerically lower or similar in Boosted compared with Unvaccinated and Primed. Significantly lower rates were reported in Boosted for Tiredness or Fatigue (22.4%), Symptoms that get worse after physical or mental activities, (1.5%), General pain/discomfort (3.0%), Difficulty breathing or shortness of breath (4.5%), Cough (0.0%), Fast-beating or pounding heart (4.5%), Headache (7.5%), Difficulty thinking or concentrating (7.5%), Sleep problems (4.5%), Mood changes (4.5%), Memory loss (3.0%) (Table 2).

### EQ-5D-5L

The study participants with long COVID (≥3 symptoms at Week 4, N=130) reported a pre-COVID mean EQ-VAS and Utility Index (UI) scores of, respectively, 84.9 and 0.879 (Table 4). Such values were not significantly different by vaccination status.

**Table 4.**
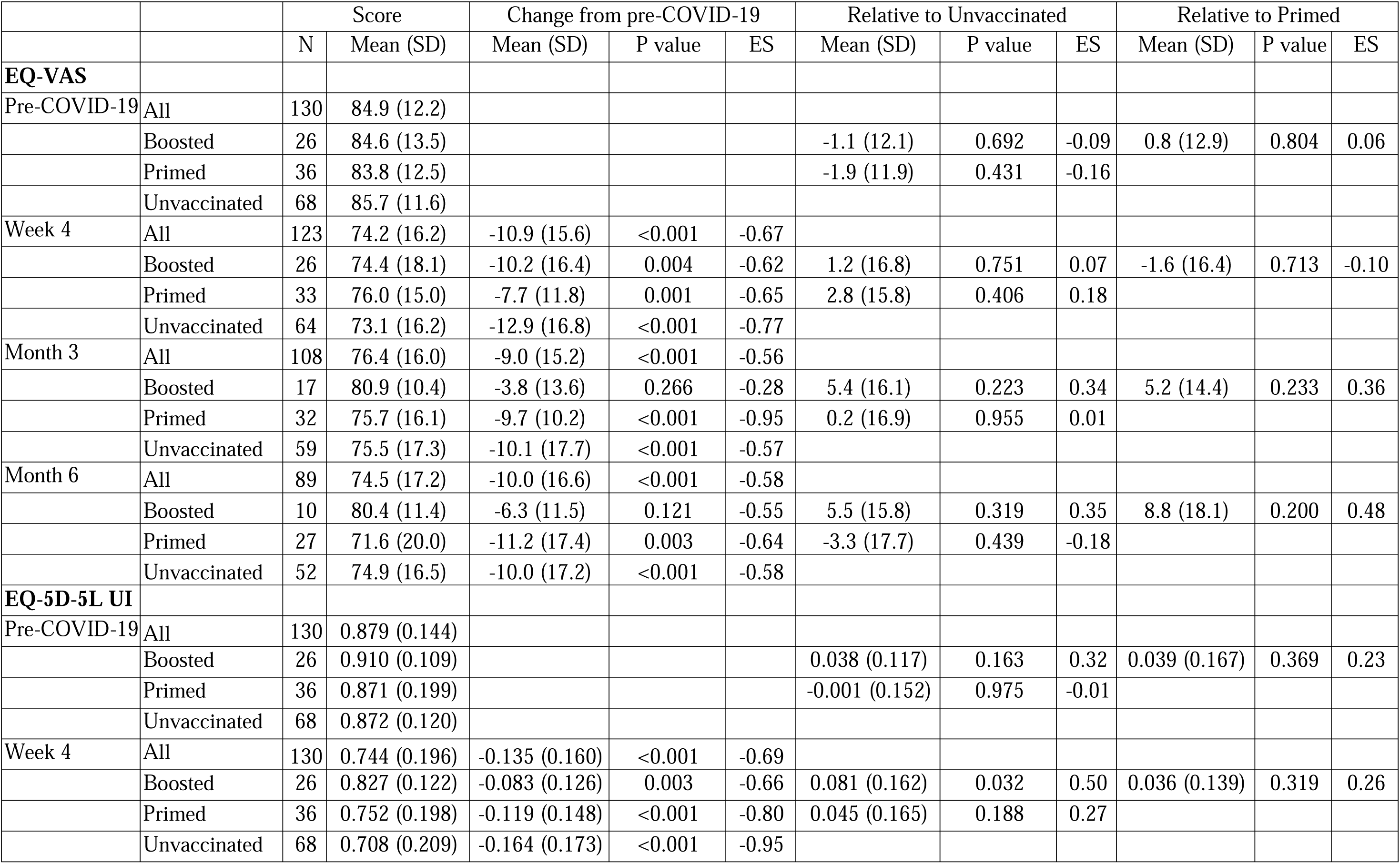
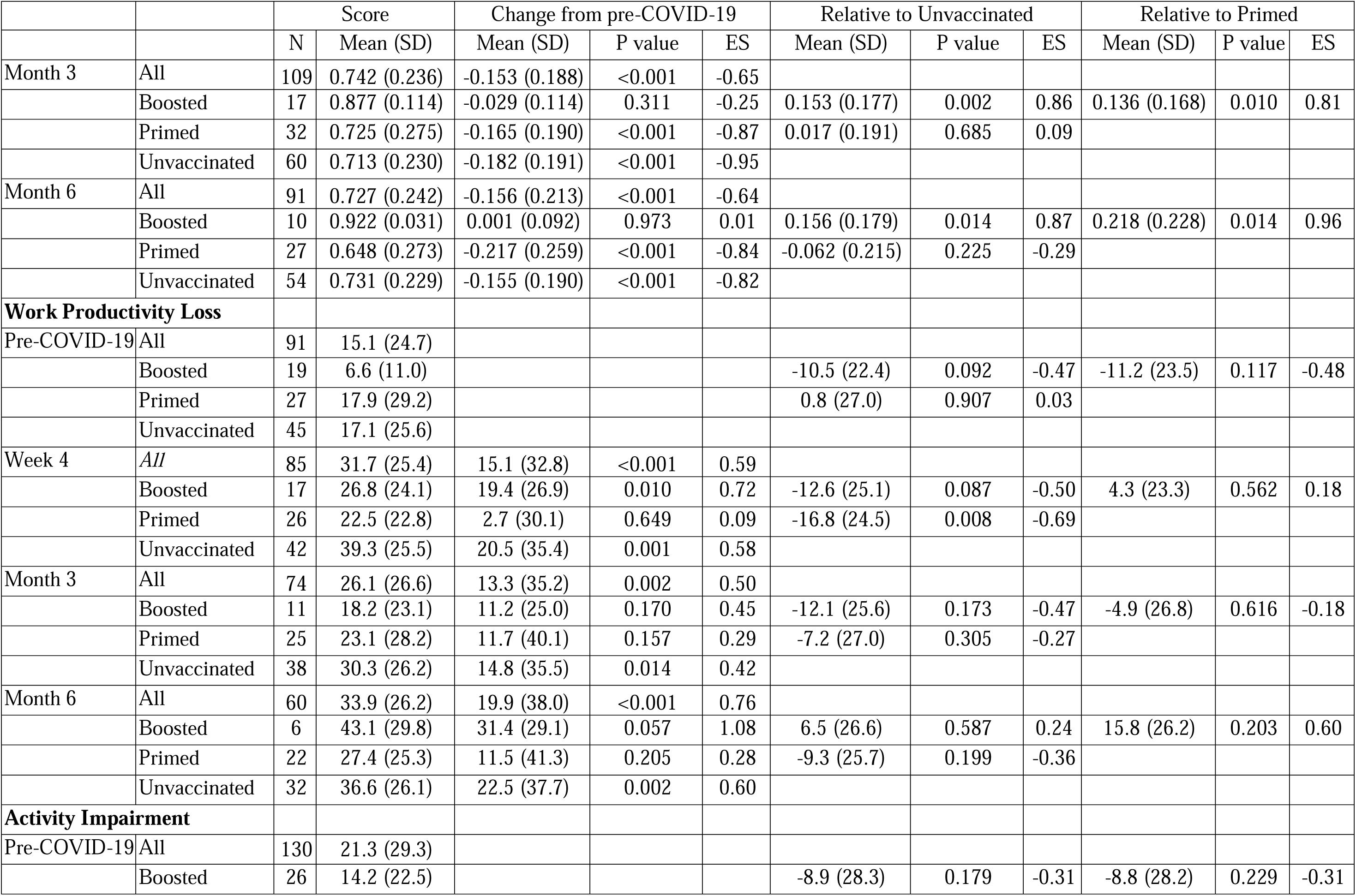
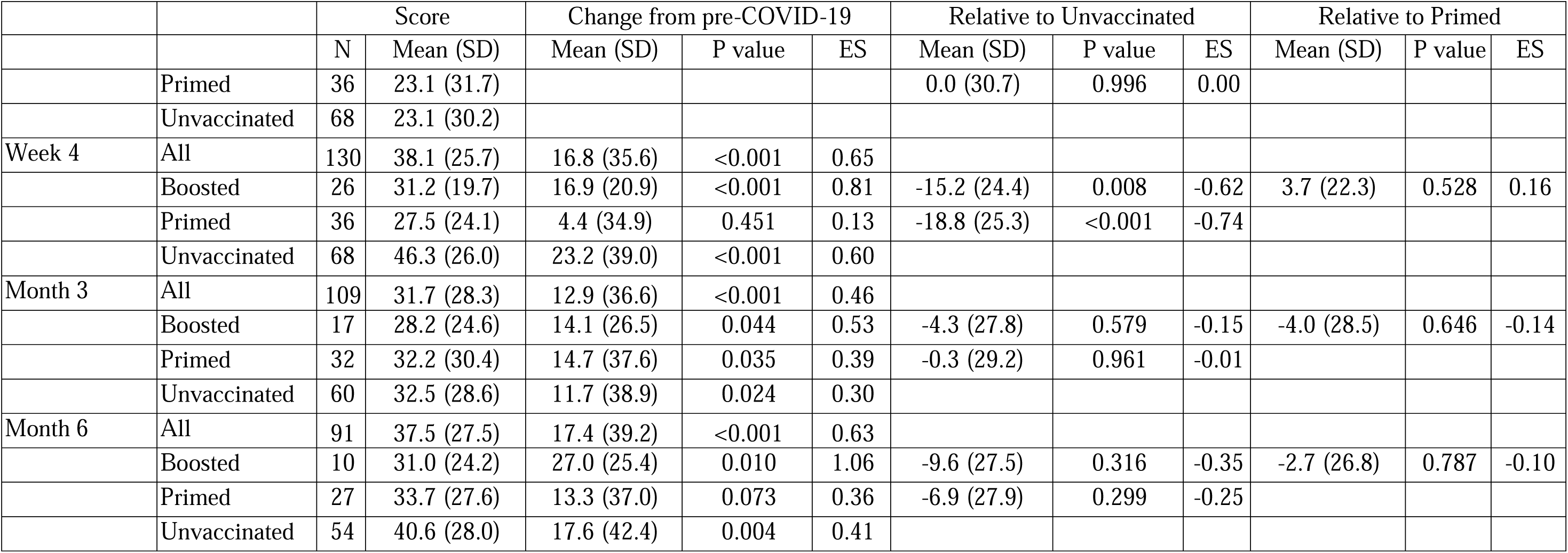
Summary of Observed EQ-5D-5L and WPAI scores by assessment time and vaccination status in long COVID subjects

|  |  | Score |  | Change from pre-COVID-19 |  |  | Relative to Unvaccinated |  |  | Relative to Primed |  |  |
| --- | --- | --- | --- | --- | --- | --- | --- | --- | --- | --- | --- | --- |
|  |  | N | Mean (SD) | Mean (SD) | P value | ES | Mean (SD) | P value | ES | Mean (SD) | P value | ES |
| **EQ-VAS** |  |  |  |  |  |  |  |  |  |  |  |  |
| Pre-COVID-19 | All | 130 | 84.9 (12.2) |  |  |  |  |  |  |  |  |  |
|  | Boosted | 26 | 84.6 (13.5) |  |  |  | -1.1 (12.1) | 0.692 | -0.09 | 0.8 (12.9) | 0.804 | 0.06 |
|  | Primed | 36 | 83.8 (12.5) |  |  |  | -1.9 (11.9) | 0.431 | -0.16 |  |  |  |
|  | Unvaccinated | 68 | 85.7 (11.6) |  |  |  |  |  |  |  |  |  |
| Week 4 | All | 123 | 74.2 (16.2) | -10.9 (15.6) | <0.001 | -0.67 |  |  |  |  |  |  |
|  | Boosted | 26 | 74.4 (18.1) | -10.2 (16.4) | 0.004 | -0.62 | 1.2 (16.8) | 0.751 | 0.07 | -1.6 (16.4) | 0.713 | -0.10 |
|  | Primed | 33 | 76.0 (15.0) | -7.7 (11.8) | 0.001 | -0.65 | 2.8 (15.8) | 0.406 | 0.18 |  |  |  |
|  | Unvaccinated | 64 | 73.1 (16.2) | -12.9 (16.8) | <0.001 | -0.77 |  |  |  |  |  |  |
| Month 3 | All | 108 | 76.4 (16.0) | -9.0 (15.2) | <0.001 | -0.56 |  |  |  |  |  |  |
|  | Boosted | 17 | 80.9 (10.4) | -3.8 (13.6) | 0.266 | -0.28 | 5.4 (16.1) | 0.223 | 0.34 | 5.2 (14.4) | 0.233 | 0.36 |
|  | Primed | 32 | 75.7 (16.1) | -9.7 (10.2) | <0.001 | -0.95 | 0.2 (16.9) | 0.955 | 0.01 |  |  |  |
|  | Unvaccinated | 59 | 75.5 (17.3) | -10.1 (17.7) | <0.001 | -0.57 |  |  |  |  |  |  |
| Month 6 | All | 89 | 74.5 (17.2) | -10.0 (16.6) | <0.001 | -0.58 |  |  |  |  |  |  |
|  | Boosted | 10 | 80.4 (11.4) | -6.3 (11.5) | 0.121 | -0.55 | 5.5 (15.8) | 0.319 | 0.35 | 8.8 (18.1) | 0.200 | 0.48 |
|  | Primed | 27 | 71.6 (20.0) | -11.2 (17.4) | 0.003 | -0.64 | -3.3 (17.7) | 0.439 | -0.18 |  |  |  |
|  | Unvaccinated | 52 | 74.9 (16.5) | -10.0 (17.2) | <0.001 | -0.58 |  |  |  |  |  |  |
| **EQ-5D-5L UI** |  |  |  |  |  |  |  |  |  |  |  |  |
| Pre-COVID-19 | All | 130 | 0.879 (0.144) |  |  |  |  |  |  |  |  |  |
|  | Boosted | 26 | 0.910 (0.109) |  |  |  | 0.038 (0.117) | 0.163 | 0.32 | 0.039 (0.167) | 0.369 | 0.23 |
|  | Primed | 36 | 0.871 (0.199) |  |  |  | -0.001 (0.152) | 0.975 | -0.01 |  |  |  |
|  | Unvaccinated | 68 | 0.872 (0.120) |  |  |  |  |  |  |  |  |  |
| Week 4 | All | 130 | 0.744 (0.196) | -0.135 (0.160) | <0.001 | -0.69 |  |  |  |  |  |  |
|  | Boosted | 26 | 0.827 (0.122) | -0.083 (0.126) | 0.003 | -0.66 | 0.081 (0.162) | 0.032 | 0.50 | 0.036 (0.139) | 0.319 | 0.26 |
|  | Primed | 36 | 0.752 (0.198) | -0.119 (0.148) | <0.001 | -0.80 | 0.045 (0.165) | 0.188 | 0.27 |  |  |  |
|  | Unvaccinated | 68 | 0.708 (0.209) | -0.164 (0.173) | <0.001 | -0.95 |  |  |  |  |  |  |
|  |  | N | Mean (SD) | Mean (SD) | P value | ES | Mean (SD) | P value | ES | Mean (SD) | P value | ES |
| Month 3 | All | 109 | 0.742 (0.236) | -0.153 (0.188) | <0.001 | -0.65 |  |  |  |  |  |  |
|  | Boosted | 17 | 0.877 (0.114) | -0.029 (0.114) | 0.311 | -0.25 | 0.153 (0.177) | 0.002 | 0.86 | 0.136 (0.168) | 0.010 | 0.81 |
|  | Primed | 32 | 0.725 (0.275) | -0.165 (0.190) | <0.001 | -0.87 | 0.017 (0.191) | 0.685 | 0.09 |  |  |  |
|  | Unvaccinated | 60 | 0.713 (0.230) | -0.182 (0.191) | <0.001 | -0.95 |  |  |  |  |  |  |
| Month 6 | All | 91 | 0.727 (0.242) | -0.156 (0.213) | <0.001 | -0.64 |  |  |  |  |  |  |
|  | Boosted | 10 | 0.922 (0.031) | 0.001 (0.092) | 0.973 | 0.01 | 0.156 (0.179) | 0.014 | 0.87 | 0.218 (0.228) | 0.014 | 0.96 |
|  | Primed | 27 | 0.648 (0.273) | -0.217 (0.259) | <0.001 | -0.84 | -0.062 (0.215) | 0.225 | -0.29 |  |  |  |
|  | Unvaccinated | 54 | 0.731 (0.229) | -0.155 (0.190) | <0.001 | -0.82 |  |  |  |  |  |  |
| **Work Productivity Loss** |  |  |  |  |  |  |  |  |  |  |  |  |
| Pre-COVID-19 | All | 91 | 15.1 (24.7) |  |  |  |  |  |  |  |  |  |
|  | Boosted | 19 | 6.6 (11.0) |  |  |  | -10.5 (22.4) | 0.092 | -0.47 | -11.2 (23.5) | 0.117 | -0.48 |
|  | Primed | 27 | 17.9 (29.2) |  |  |  | 0.8 (27.0) | 0.907 | 0.03 |  |  |  |
|  | Unvaccinated | 45 | 17.1 (25.6) |  |  |  |  |  |  |  |  |  |
| Week 4 | All | 85 | 31.7 (25.4) | 15.1 (32.8) | <0.001 | 0.59 |  |  |  |  |  |  |
|  | Boosted | 17 | 26.8 (24.1) | 19.4 (26.9) | 0.010 | 0.72 | -12.6 (25.1) | 0.087 | -0.50 | 4.3 (23.3) | 0.562 | 0.18 |
|  | Primed | 26 | 22.5 (22.8) | 2.7 (30.1) | 0.649 | 0.09 | -16.8 (24.5) | 0.008 | -0.69 |  |  |  |
|  | Unvaccinated | 42 | 39.3 (25.5) | 20.5 (35.4) | 0.001 | 0.58 |  |  |  |  |  |  |
| Month 3 | All | 74 | 26.1 (26.6) | 13.3 (35.2) | 0.002 | 0.50 |  |  |  |  |  |  |
|  | Boosted | 11 | 18.2 (23.1) | 11.2 (25.0) | 0.170 | 0.45 | -12.1 (25.6) | 0.173 | -0.47 | -4.9 (26.8) | 0.616 | -0.18 |
|  | Primed | 25 | 23.1 (28.2) | 11.7 (40.1) | 0.157 | 0.29 | -7.2 (27.0) | 0.305 | -0.27 |  |  |  |
|  | Unvaccinated | 38 | 30.3 (26.2) | 14.8 (35.5) | 0.014 | 0.42 |  |  |  |  |  |  |
| Month 6 | All | 60 | 33.9 (26.2) | 19.9 (38.0) | <0.001 | 0.76 |  |  |  |  |  |  |
|  | Boosted | 6 | 43.1 (29.8) | 31.4 (29.1) | 0.057 | 1.08 | 6.5 (26.6) | 0.587 | 0.24 | 15.8 (26.2) | 0.203 | 0.60 |
|  | Primed | 22 | 27.4 (25.3) | 11.5 (41.3) | 0.205 | 0.28 | -9.3 (25.7) | 0.199 | -0.36 |  |  |  |
|  | Unvaccinated | 32 | 36.6 (26.1) | 22.5 (37.7) | 0.002 | 0.60 |  |  |  |  |  |  |
| **Activity Impairment** |  |  |  |  |  |  |  |  |  |  |  |  |
| Pre-COVID-19 | All | 130 | 21.3 (29.3) |  |  |  |  |  |  |  |  |  |
|  | Boosted | 26 | 14.2 (22.5) |  |  |  | -8.9 (28.3) | 0.179 | -0.31 | -8.8 (28.2) | 0.229 | -0.31 |
|  |  | N | Mean (SD) | Mean (SD) | P value | ES | Mean (SD) | P value | ES | Mean (SD) | P value | ES |
|  | Primed | 36 | 23.1 (31.7) |  |  |  | 0.0 (30.7) | 0.996 | 0.00 |  |  |  |
|  | Unvaccinated | 68 | 23.1 (30.2) |  |  |  |  |  |  |  |  |  |
| Week 4 | All | 130 | 38.1 (25.7) | 16.8 (35.6) | <0.001 | 0.65 |  |  |  |  |  |  |
|  | Boosted | 26 | 31.2 (19.7) | 16.9 (20.9) | <0.001 | 0.81 | -15.2 (24.4) | 0.008 | -0.62 | 3.7 (22.3) | 0.528 | 0.16 |
|  | Primed | 36 | 27.5 (24.1) | 4.4 (34.9) | 0.451 | 0.13 | -18.8 (25.3) | <0.001 | -0.74 |  |  |  |
|  | Unvaccinated | 68 | 46.3 (26.0) | 23.2 (39.0) | <0.001 | 0.60 |  |  |  |  |  |  |
| Month 3 | All | 109 | 31.7 (28.3) | 12.9 (36.6) | <0.001 | 0.46 |  |  |  |  |  |  |
|  | Boosted | 17 | 28.2 (24.6) | 14.1 (26.5) | 0.044 | 0.53 | -4.3 (27.8) | 0.579 | -0.15 | -4.0 (28.5) | 0.646 | -0.14 |
|  | Primed | 32 | 32.2 (30.4) | 14.7 (37.6) | 0.035 | 0.39 | -0.3 (29.2) | 0.961 | -0.01 |  |  |  |
|  | Unvaccinated | 60 | 32.5 (28.6) | 11.7 (38.9) | 0.024 | 0.30 |  |  |  |  |  |  |
| Month 6 | All | 91 | 37.5 (27.5) | 17.4 (39.2) | <0.001 | 0.63 |  |  |  |  |  |  |
|  | Boosted | 10 | 31.0 (24.2) | 27.0 (25.4) | 0.010 | 1.06 | -9.6 (27.5) | 0.316 | -0.35 | -2.7 (26.8) | 0.787 | -0.10 |
|  | Primed | 27 | 33.7 (27.6) | 13.3 (37.0) | 0.073 | 0.36 | -6.9 (27.9) | 0.299 | -0.25 |  |  |  |
|  | Unvaccinated | 54 | 40.6 (28.0) | 17.6 (42.4) | 0.004 | 0.41 |  |  |  |  |  |  |

At Week 4, the observed (Table 4) and model-based (Table 5) EQ-VAS scores were lower than pre-COVID, regardless of vaccination status (not significantly for Primed according to the model-based estimates). The observed EQ-VAS scores for the entire long COVID population were numerically similar between Week 4 (74.2) and Month 6 (74.5), and, by the end of the study period, they did not return to pre-COVID levels. The observed EQ-VAS scores in Unvaccinated and Primed were still significantly lower than pre-COVID at Month 3 and 6; only Boosted reported EQ-VAS levels not different than pre-COVID at Month 3 (p=0.266) and Month 6 (p=0.121). The model-based EQ-VAS scores were numerically lower at Month 3 and 6 regardless of vaccination status, while not significantly different from pre-COVID, except for Primed at Month 6 (Table 5).

**Table 5.**
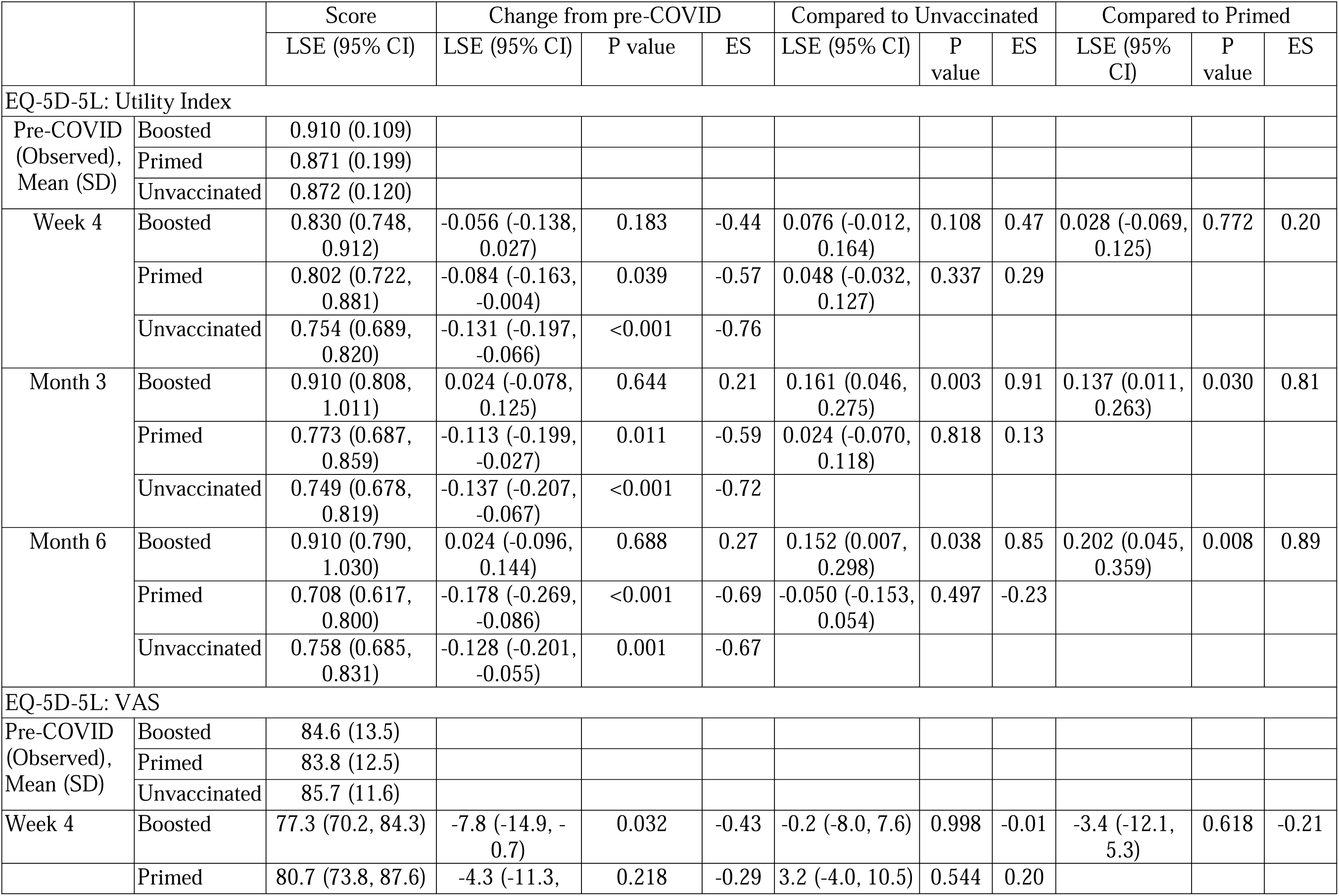
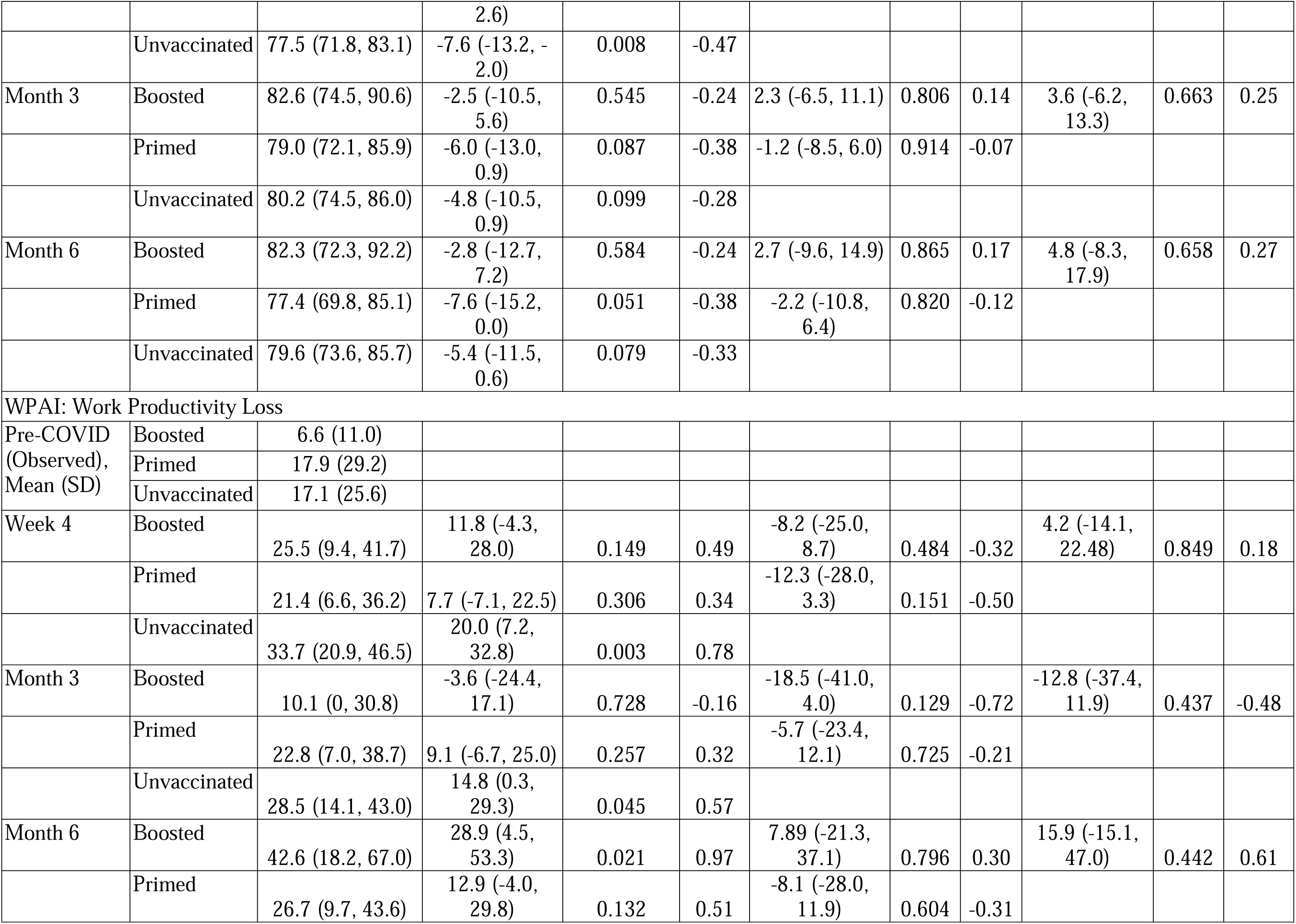
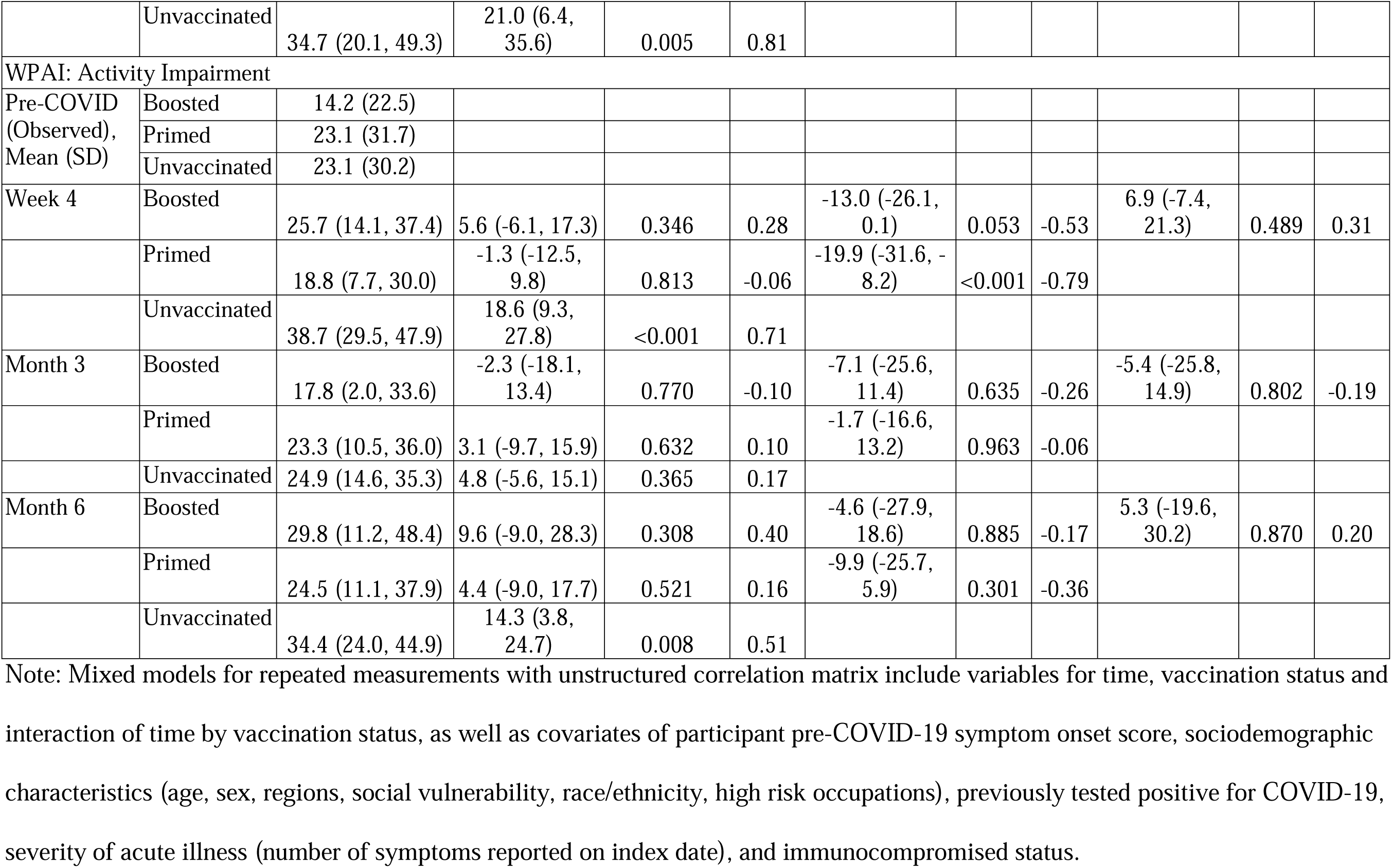
Least-Square Mean Estimates and 95% CI of EQ-5D-5L and WPAI-GH scores in long COVID subjects

Both observed (Table 4) and model-based (Table 5) UI scores were lower than pre-COVID for Unvaccinated and Primed for Month 3 and 6 while not different among Boosted. Consistently, the model-based UI scores of Boosted were not different from pre-COVID at Month 3, 6 and since Week 4, too (Table 5).

The observed UIs values for the entire long-COVID population were numerically similar between Week 4 (0.74), Month 3 (0.74) and 6 (0.73) and did not return to pre-COVID levels. Such effect was driven by the detrimental effect of COVID-19 reported by Unvaccinated and Primed. At Week 4, the mean UI change from pre-COVID in Boosted was significantly lower than in Unvaccinated based on observed data and numerically lower according to model-based estimates.

The mean changes in UIs from pre-COVID in Boosted were significantly lower versus Unvaccinated at Month 3 and 6 based on both observed and model estimates, with medium-to-high effect sizes. The mean changes from pre-COVID in UIs reported by Boosted were significantly lower than Primed at Month 3 and 6 based on both observed and model estimates, with medium-to-high effect sizes.

The impact of COVID-19 on the HRQoL was detrimental for participants with long COVID, but significantly less so for Boosted, the group with the highest UI and EQ-VAS scores during the 6-month study period.

### WPAI

The WPAI analyses had a smaller eligible population and were impacted by small sample size. A total of 85 long COVID study participants (66%) reported being currently employed at Week 4 and were eligible to complete the work productivity questions. Of those, 42 (49%) Unvaccinated, 26 (31%) Primed and 17 (20%) Boosted.

Participants reported a pre-COVID mean Work Productivity (WP) loss of 15.1% (Table 4). Such values were not significantly different by vaccination status. At Week 4, the observed WP loss increased substantially to 31.7%. The observed WP loss scores for the entire long-COVID population were numerically similar between Week 4 (31.7%), Month 3 (26.1%) and Month 6 (33.9%), and did not return to pre-COVID levels. Such effect was driven by the detrimental effect of COVID-19 reported by Unvaccinated. At Week 4, both observed and model-based WP scores were significantly lower than pre-COVID for the Unvaccinated. At Month 3 and 6, the observed and model-based mean change in WP levels were not significantly different than pre-COVID for Boosted and Primed (non-significant for model-based estimated for Boosted at Month 6), suggesting a return to pre-COVID levels for these vaccinated groups only.

The long COVID study participants (N=130) reported a pre-COVID mean Activity Impairment (AI) of 21.3% (Table 4). Such values were not significantly different by vaccination status. At Week 4, the observed AI increased substantially to 38.1%. The observed AI scores for the entire long-COVID population were numerically similar between Week 4 (38.1%), Month 3 (31.7%) and Month 6 (37.5%), and did not return to pre-COVID levels. Such effect was driven by the detrimental effect of COVID-19 reported by Unvaccinated, who reported the highest AI across all time points. At Week 4, the mean change from pre-COVID was significantly lower than Unvaccinated for Boosted and Primed based on both the observed and model-based estimates. At Month 3 and Month 6, all groups were still highly impacted; subjects Boosted reported the lowest AI levels.

## Discussion

In this national observational study conducted among US symptomatic outpatients with a documented SARS-CoV-2 infection, long COVID had a detrimental effect on well-being, work productivity and activity levels. Long COVID symptoms were found persistent over 6 months in over a third of the study cohort, resulting in prolonged limitation of activities and work productivity. Compared with Primed and Unvaccinated, subjects Boosted with BNT162b2 prior to a breakthrough infection were associated with significantly lower likelihood of long COVID onset, fewer symptoms, and faster improvement over time. Positive trends in the outcomes assessed were observed for Primed versus Unvaccinated, although generally not statistically significant, most likely due to the relatively long mean time since last dose for Primed. The prevalence of enduring symptoms after a mild COVID infection in this study was generally consistent with published literature[1–5, 8]. The protective association was consistent based on observed and model-based analyses and using alternative definitions of long COVID. These findings support growing research and consensus that prior COVID-19 vaccination may have a protective effect against long COVID [4–8].

This study has several strengths compared with prior research. From a novelty perspective, this study is one of a limited number assessing diverse PROs associated with long COVID in a nationwide real-world source population. As such, it contributes a holistic picture of the long-term humanistic outcomes of COVID-19, assessed directly from a patient’s perspective. From an internal validity perspective, the study enrolled patients within days from testing positive and prospectively collected survey-based data, potentially reducing recall bias risks. Moreover, the study leveraged widely used validated PRO instruments (EQ-5D-5L, WPAI) and a questionnaire capturing a comprehensive symptoms list aligned to CDC research on long COVID [1]. With asymptomatic infections excluded by design, our estimates can be interpreted exclusively as related to symptomatic disease, potentially reducing the risk of overestimating the prevalence of long COVID among symptomatic. Further, both observed and model-based analyses yielded consistent results, and our findings were robust using alternative case definitions. Finally, with all study activities carried out virtually, this study piloted an innovative approach to agile and digitally enabled research during a pandemic.

The study is subject to several limitations. As previously described [9], all data collected was self-reported, subject to missingness, errors, recall bias, social desirability bias and selection bias associated with survey drop-off. Out of 328 participants, 21% dropped off at Month 6, possibly due to response fatigue and/or survey burden. Such drop-off rate should be interpreted in the context of participants being asked not to skip surveys. Such a strict requirement allowed for a clean assessment of changes in outcomes prevalence over time, but at the cost of attrition. Other limitations included the female over-representation, the relatively healthy status of the source population, and the fact that the study focused on adults only [9]. Further, the long COVID definitions used were based on presence of symptoms, with no assessment of severity of symptoms. Despite adjusting for several covariates, risk of residual confounding remains. Finally, these findings may not be generalizable to prior or future variants, other countries, time periods and populations that were excluded.

Booster vaccination with BNT162b2 has been shown to be safe and effective at reducing the risk of infection and potentially protective against long COVID [6, 8]; this study shows its potential beneficial effect in preventing long COVID and attenuating its burden. These outcomes can inform estimation of quality-adjusted life years and indirect costs. While this study contributes to addressing knowledge gaps related to long COVID, the characterization of long COVID continues to evolve [28–30]. Future studies could corroborate these findings with different data collection methods and designs, considering a non-COVID comparator, longer follow-up times, use of COVID-19 specific validated instruments, as well as if these protective effects may minimize impact on other individuals and households.

## Conclusions

Long COVID had a detrimental effect on well-being, work productivity and activity levels. Booster vaccination with BNT162b2 generally offered the highest level of protection against long COVID. The booster was associated with less symptomatic infection and faster improvement, with boosted subjects experiencing the lowest number of symptoms over time. This, in turn, contributed to improved HRQoL and maintained productivity and activities. These findings support current recommendations for broad use of BNT162b2.

## Data Availability

Aggregated data that support the findings of this study are available upon reasonable request from the corresponding author MDF, subject to review. These data are not publicly available due to them containing information that could compromise research participant privacy/consent.

## Abbreviations

ANOVA: Analysis of variance
CDC: Centers for Disease Control and Prevention
CI: confidence interval; ES: effect size
SD: standard deviation
SVI: social vulnerability index
UI: utility index
VAS: visual analogue scale
WPAI: work productivity and impairment

## Declarations

### Ethics approval and consent to participate

This study was approved by the Sterling IRB, Protocol #C4591034. Participation in the study was voluntary and anonymous. Consent was obtained electronically via the CVS Health E-Consent platform. Participants were informed of their right to refuse or withdraw from the study at any time. Participants were compensated for their time.

### Consent for publication

All authors have given their approval for this manuscript version to be published.

### Author contributions

All named authors meet the International Committee of Medical Journal Editors (ICMJE) criteria for authorship for this article. XS: Conceptualization, Methodology, Data curation, Formal analysis, Writing - original draft. MDF and YP: Investigation, Project administration, Conceptualization, Writing - Review & editing. MMM, HC, LP, MBA, and JMZ: Conceptualization, Writing - Review & editing. JCC: Conceptualization, Methodology, Formal analysis, Writing - Review & editing.

## Acknowledgements

The authors acknowledge Nancy Gifford (Pfizer employee), Joseph Ferenchick, Shiyu Lin and Shawn Edmonds (CVS Health employees) for specific contributions to this research project. Editorial support was provided by Laura Anatale-Tardiff and Leena Samuel at CVS Health and was funded by Pfizer.

## Supplemental Material

**Supplemental Figure 1.**
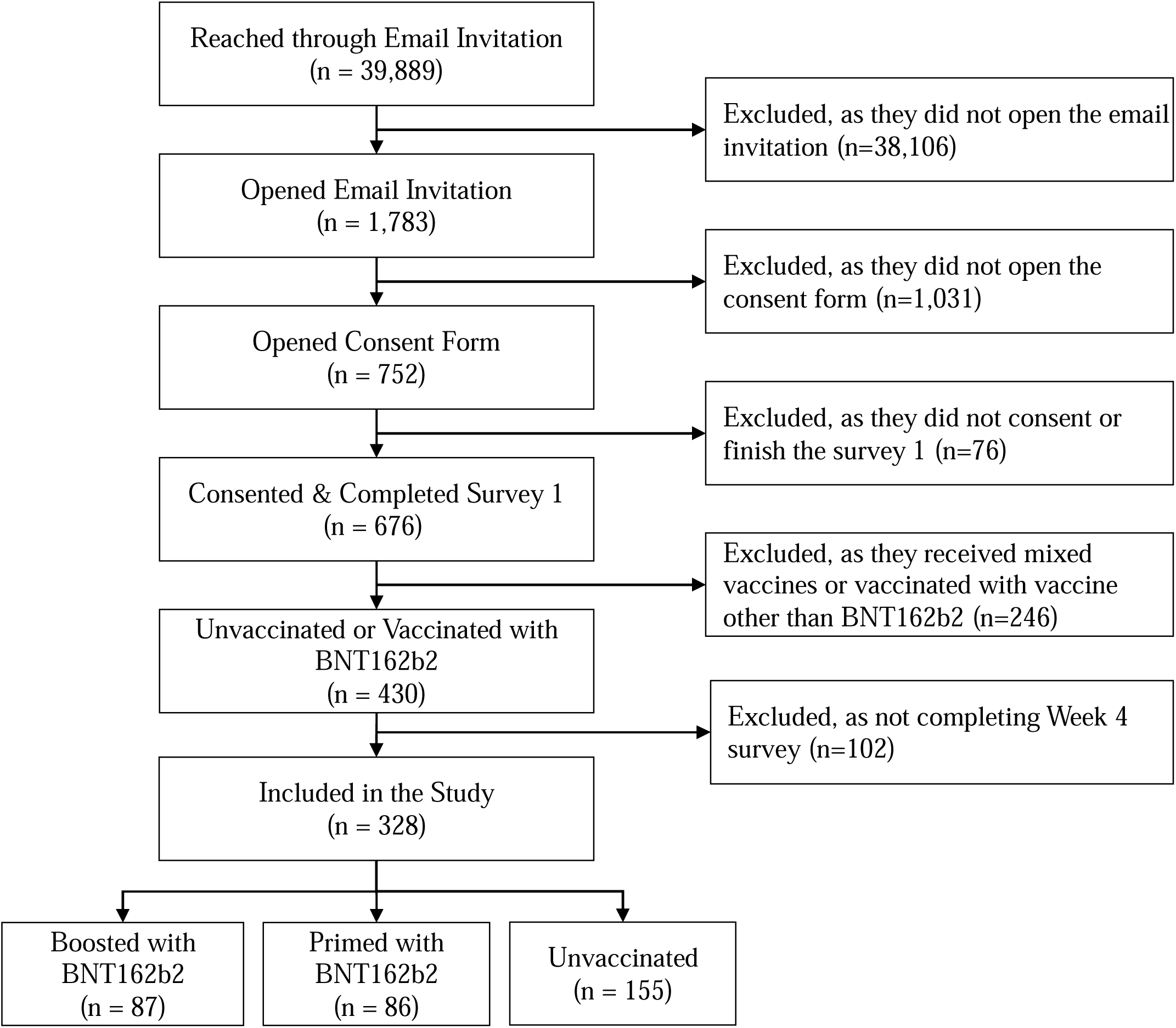
Study Flow Chart

**Supplemental Figure 2.**
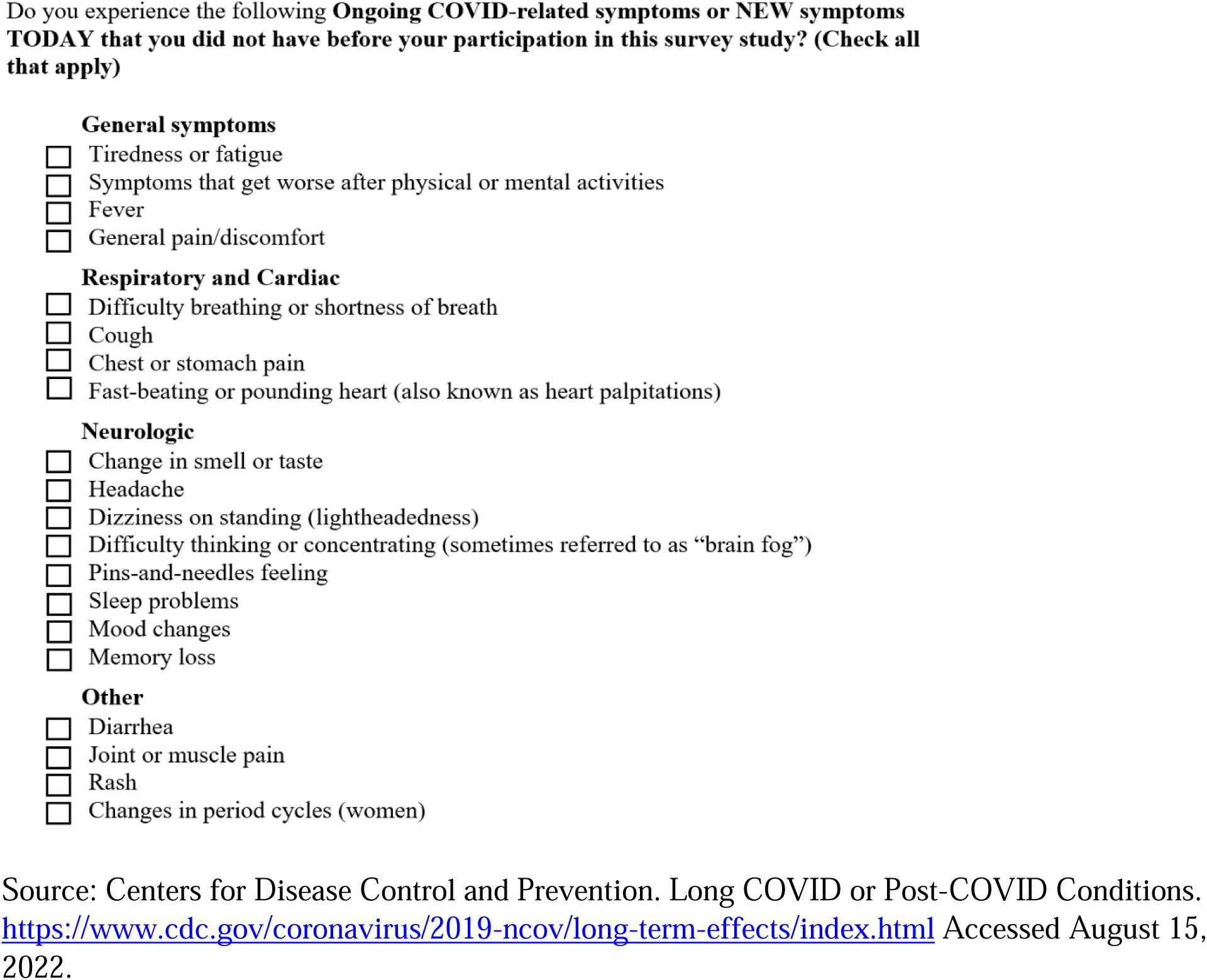
Questionnaire on long COVID symptoms

**Supplemental Figure 3.**
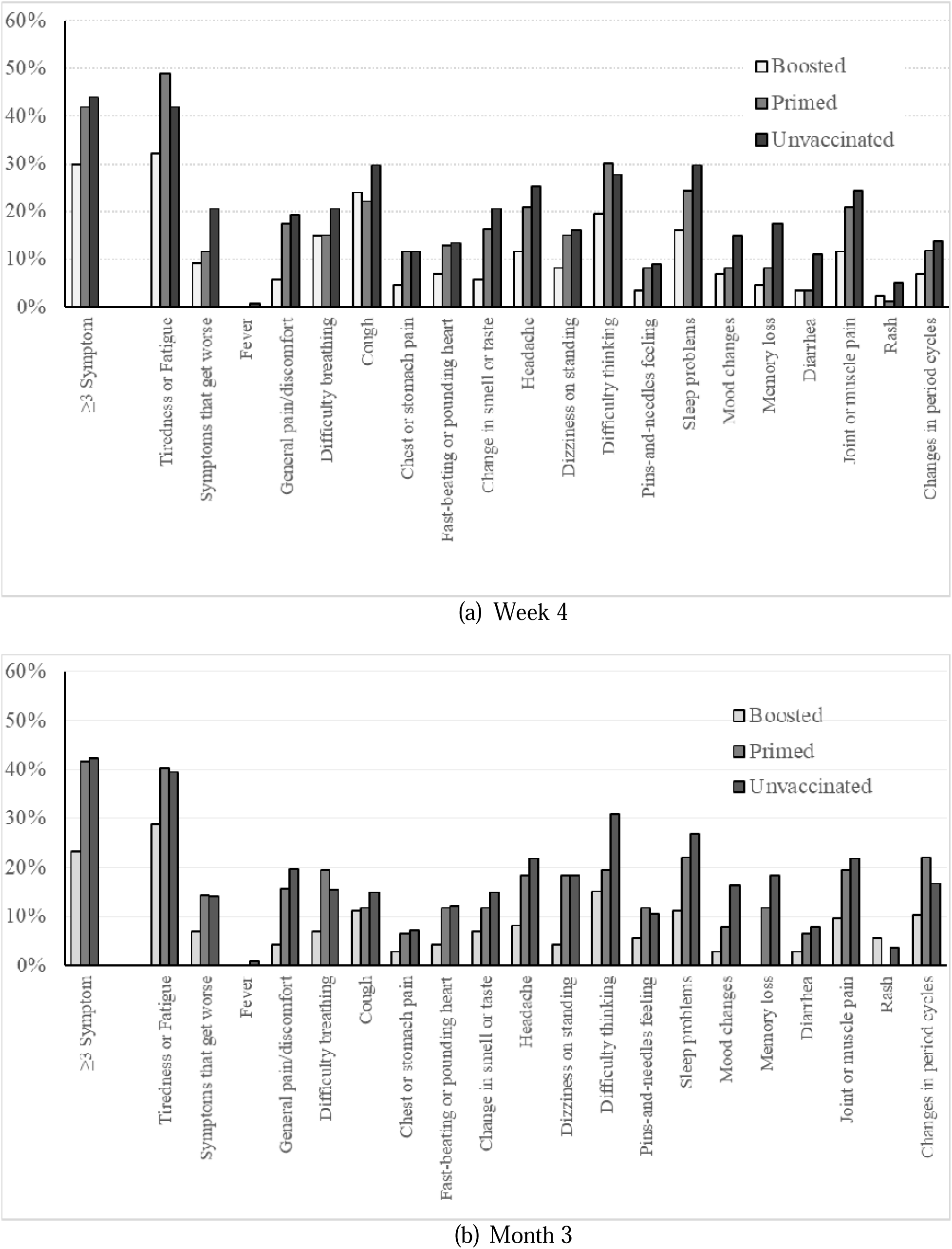
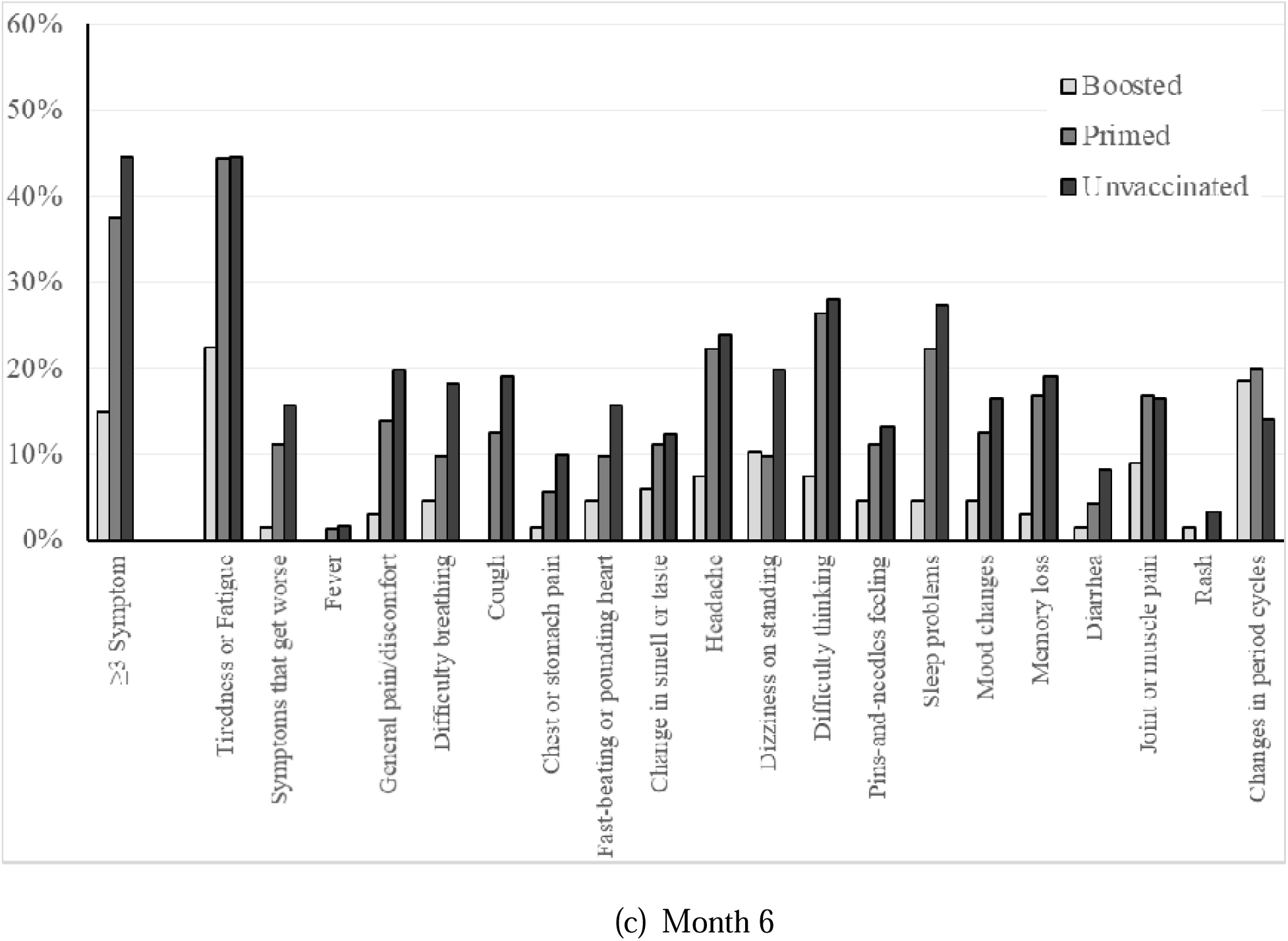
Prevalence of long COVID symptoms by vaccination status

**Supplemental Figure 4.**
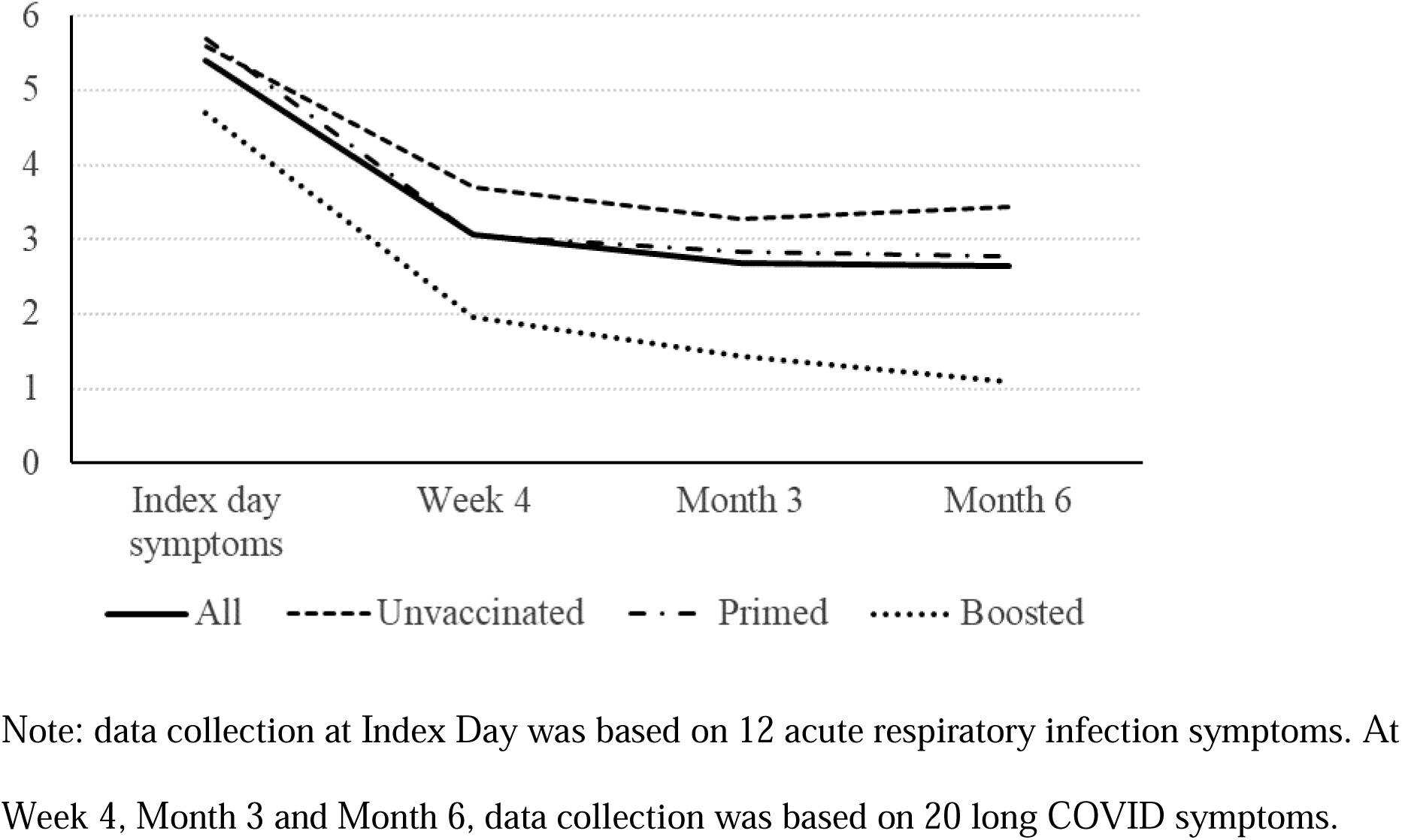
Number of symptoms over time by vaccination status

**Supplemental Figure 5.**
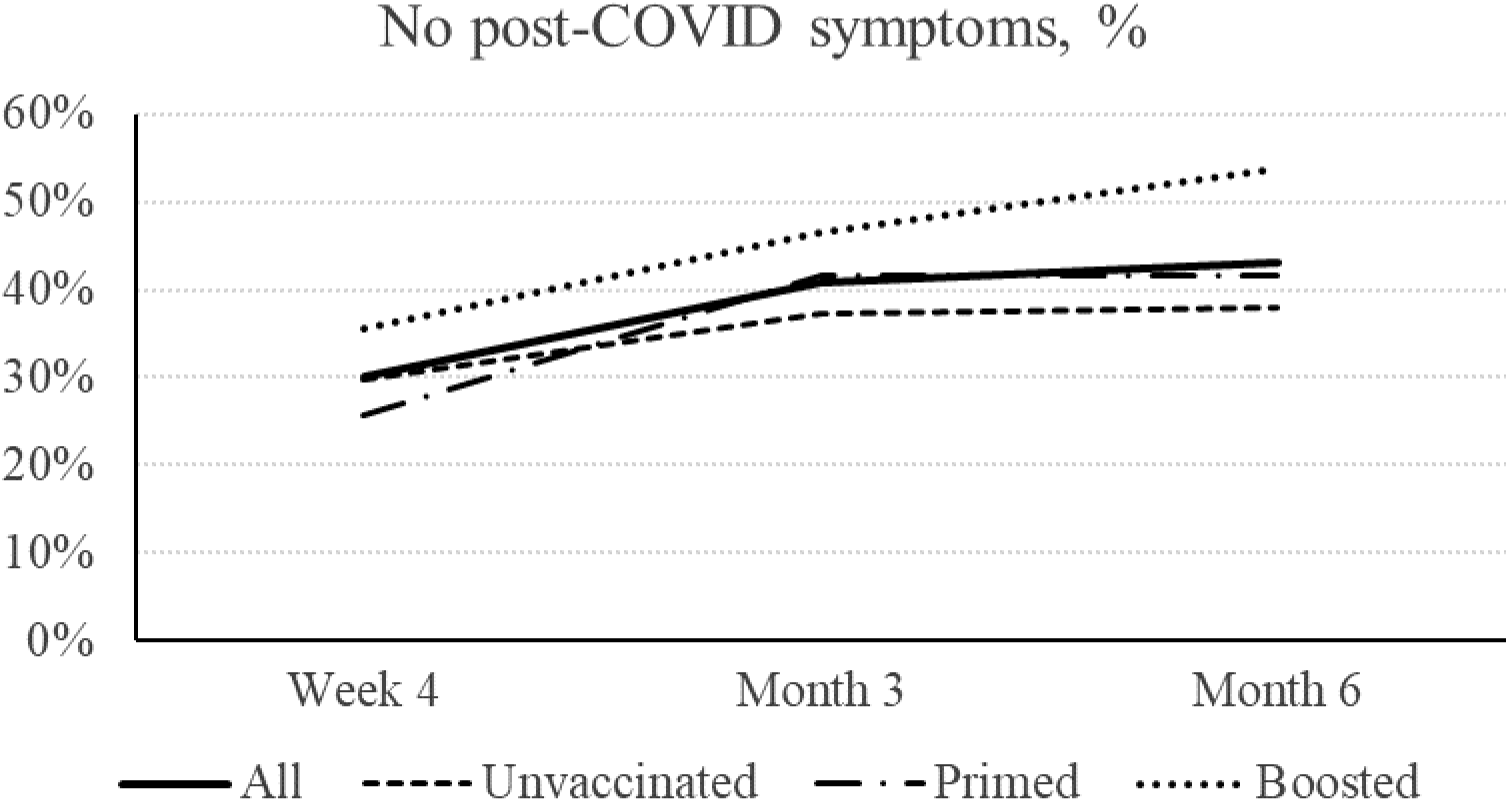
Absence of long COVID symptoms by vaccination status

